# Family Constellations for All Clinical Conditions: A Systematic Review and Meta-analysis Showing a Lack of Supporting Evidence

**DOI:** 10.64898/2026.04.19.26351231

**Authors:** Filipe Luis Souza, Nathalia Cabral Souza, Josimar Antônio de Alcântara Mendes

## Abstract

**Introduction:** Family Constellation Therapy (FCT) has been widely disseminated in clinical, public health, and judicial settings despite persistent concerns regarding its theoretical basis, safety, and the limited availability of rigorous randomised evidence supporting its clinical use.

**Objective:** The aim of this systematic review is to assess the effects of FCT across all clinical conditions, explicitly considering both benefits and harms; and summarise the characteristics of studies and intervention settings used in randomised controlled trials of FCT.

**Methods:** Following a prospectively registered protocol (CRD420251136190), we conducted a systematic search of seven databases (PubMed, EMBASE, APA PsycInfo, CENTRAL, BVS, Web of Science, and CINAHL) and grey literature (ICTRP and ProQuest database) without language or date restrictions to identify published and unpublished randomised controlled trials of FCT. Study selection, data extraction, risk of bias (RoB 2), and certainty of evidence (GRADE) were performed in duplicate. Statistical analyses followed a prospectively registered analysis plan with prespecified criteria for data pooling and for handling analytical limitations.

**Results:** No reliable evidence was found to support the use of FCT for any condition across both clinical and non-clinical samples. All trials included were judged to be at high risk of bias and all comparisons were rated as very low-certainty evidence. Concerns regarding potential adverse effects were identified, and the available data was insufficient to establish the effectiveness of the intervention, precluding any clinical recommendation.

**Conclusion:** Clinicians, policymakers, and consumers should reconsider adopting FCT while reliable evidence is not available.

## 1. INTRODUCTION

Family Constellations Therapy (FCT), developed by the German practitioner Bert Hellinger in the early 1990s, is typically presented as a brief, facilitator-led group intervention. Participants and “representatives” are spatially arranged to externalise the client’s “inner image” of a conflictual social system (most often the family) and to move towards a “solution constellation” through repositioning and ritualised sentences aligned with putative systemic “orders” governing relationships.^1,2^ In Hellinger’s framing, relational dynamics are governed by universal laws of belonging, balance between giving and receiving, and hierarchy; transgressions of these “laws” purportedly produce symptoms that may reappear across generations until systemic recognition and re-ordering occur.^3,6^ In practice, this framing invites clinical, therapeutic and institutional decision-making to be anchored in claims of universality and intergenerational causation, underpinned by a strongly mystical foundation, that are presented as self-evident within the method itself, precisely where psychology and psychotherapy ordinarily require explicit theorisation, testable mechanisms and accountable standards of care.^5,6^

In recent years, multiple scholarly appraisals have characterised FCT as a pseudoscientific practice lacking a coherent evidential and theoretical basis, particularly within psychology and psychotherapy.^4,5,7,8^ Despite its growing reach in Europe and South America and its adoption by thousands of practitioners, empirical scrutiny has lagged behind dissemination, with the available literature characterised by limited randomised evidence and persistent methodological.^4,5,8,9^ This mismatch between expansive claims and thin evidence is not a merely academic concern, it has direct ethical and safeguarding implications: concerns about potential harms prompted by FCT have been raised on ethical and clinical grounds, including the possibility of precipitating acute distress or disorganisation without appropriate safeguards, operating in tension with duties of evidence-based care and confidentiality, and exposing clients to high-intensity interventions without the procedural protections that are routine in regulated psychotherapeutic practice.^4,5,7,8^ Some scholars argue that FCT does not meet core psychotherapy standards regarding process, contracting, diagnosis, supervision, confidentiality and post-intervention care, cautioning against its provision in professional settings, a conclusion that renders continued professional endorsement difficult to defend on therapeutic and safeguarding grounds.^6,7,8,10^ Others note that these risks become especially severe in contexts marked by domestic violence, where FCT practices may facilitate ethically problematic dynamics and elevate the risk of revictimisation, precisely when safeguarding should be most robust.^3,5^

These risks and evidentiary uncertainties are particularly salient where FCT has been incorporated into public health and judicial settings, a scenario that intensifies the ethical stakes by normalising a practice that remains contested on scientific and clinical grounds. In Brazil, for example, debates have highlighted its diffusion into courts and public services, including critiques of funding through the Unified Health System (SUS), raising questions about scientific legitimacy, safeguarding and human rights implications.^3,4,5^ Brazilian national professional guidance further warns psychologists that FCT does not constitute a recognised psychological method or technique and flags ethical incompatibilities with professional practice.^10^ Against this backdrop, there is a clear need for a rigorous, transparent synthesis focused specifically on the potential clinical effects and safety of FCT. Accordingly, this systematic review will: a) assess the effects of FCT across all clinical conditions, explicitly considering both benefits and harms; and b) summarise the characteristics of studies and intervention settings used in randomised controlled trials of FCT.

## 2. METHODS

This systematic review was conducted following the guidance of the *Cochrane Handbook for Systematic Reviews of Interventions* (version 6.5)^11^ and reported in accordance with the Preferred Reporting Items for Systematic Reviews and Meta-Analyses (PRISMA) 2020 statement^12^ and was preregistered in PROSPERO (CRD420251136190) prior to the conduct of any formal searches or analyses. A systematic literature search was undertaken across 7 electronic databases: MEDLINE (via PubMed), EMBASE (via Ovid), APA PsycInfo (via Ovid), CENTRAL, Biblioteca Virtual em Saúde (BVS; including records in English, Brazilian Portuguese, and Spanish), Web of Science, and CINAHL. To identify grey literature and capture potentially unpublished or ongoing studies with available results, additional records were retrieved from the International Clinical Trials Registry Platform (ICTRP), the ProQuest database, and through manual handsearching the reference list of included studies, previous literature reviews on the topic, and contacting experts about ongoing research. No restrictions were applied regarding year of publication or language of dissemination. Searches were conducted retrospectively and covered all records available up to 27 November 2025. Eligibility criteria were defined using the Population, Intervention, Comparison, Outcome, Timing, and Study Design (PICOTS) framework in a comprehensive manner to include all published and unpublished randomised controlled trials (RCTs) on FCT. Detailed search strategies and database-specific adaptations are provided in Appendices 1, 2, 3 and 4.

### 2.1. Study selection

Study selection was conducted using Covidence.^13^ Following automated (Covidence) deduplication, two reviewers (F.L.S. and N.C.S.) independently and in duplicate screened each record by title and abstract, followed by screening through full-text reading. Prior to formal screening, both reviewers completed a joint pilot exercise by screening ten randomly selected records to ensure consistent interpretation of the inclusion criteria and familiarity with the platform. Disagreements were resolved by consultation with a third reviewer (J.A.A.M.).

### 2.2. Data extraction and management

Data extraction was performed independently and in duplicate by two reviewers (F.L.S. and N.C.S.) using a standardised Microsoft Excel form developed specifically for this review. Prior to full data extraction, the reviewers completed a joint training session and piloted the extraction form using a randomised controlled trial unrelated to the review topic.

Extracted data included study characteristics, participant demographics, intervention and comparator details, outcome measures, assessment time points, and main findings. When multiple reports originated from the same trial, data were collated and treated as a single study. Any discrepancies in extracted data were resolved by consensus or, when required, by consultation with a third reviewer (J.A.A.M.).

### 2.3. Risk of Bias and Certainty of Evidence

Two reviewers (F.L.S. and N.C.S.) independently assessed the risk of bias using the Cochrane Risk of Bias 2 (RoB 2) tool.^14^ This instrument evaluates bias across predefined domains related to the randomisation process, deviations from intended interventions, missing outcome data, outcome measurement, and selective reporting. Disagreements between reviewers were resolved through discussion or, if necessary, adjudication by a third reviewer (J.A.A.M.). The certainty of the evidence for each clinical question identified in the review was determined using the Grading of Recommendations, Assessment, Development, and Evaluations (GRADE) approach,^15,16^ considering risk of bias, inconsistency, indirectness, imprecision, and publication bias. Summary of Findings tables were generated using GRADEpro tool (https://www.gradepro.org/).

The GRADE assessment was conducted independently by two reviewers (F.L.S. and N.C.S.), following the procedures pre-specified in the review protocol to ensure transparency and methodological consistency. Any discrepancies in certainty judgments were resolved through discussion, with consultation from a third reviewer (J.A.A.M.) when consensus could not be reached. To minimise post-hoc decision-making, explicit, pre-registered criteria were applied across all clinical questions to guide downgrading decisions (CRD420251136190). All judgements regarding downgrading or upgrading the certainty of evidence were documented and justified.

### 2.4. Treatment effect measures

Where appropriate, continuous data were analysed in random-effects meta-analysis using standardised mean differences (SMDs) with 95% CIs. Means and standard deviations were extracted from each study arm, and standard deviations were derived from alternative summary statistics when not directly reported. When required, outcome directions were harmonised so that higher scores consistently reflected the same construct across studies.

Meta-analyses were conducted only when at least two clinically comparable studies were available according to predefined PICOTS criteria; otherwise, results were synthesised narratively. All analyses were performed using Review Manager (RevMan) version 5.4.1. In instances where it was not possible to perform a meta-analysis for a particular PICO element, a descriptive forest plot was performed to visually display the effect estimate from the available study.

Analyses for dichotomous data were also planned, but no studies with this type of data were included. The data synthesis methods used for this review is described in detail in the preregistered protocol available online (CRD420251136190).

## 3. RESULTS

The database search identified 288 records, with 3 additional records retrieved through handsearching, yielding a total of 291 references. After the removal of 129 duplicate records identified through Covidence, 162 unique records were screened by title/abstract and assessed 5 full-text articles eligibility. No studies were excluded at the full-text stage. Consequently, five randomised controlled trials met the inclusion criteria and were included in the study (Figure 1). Of these, one trial record (NCT06863948) was identified as completed. We requested data for our review multiple times through the e-mail contact provided in the public record, and up to the time of publication of this review, we have not received a response. All studies excluded were documented, together with the specific reasons for exclusion, and provided in Appendices 6 and 7. Four trials provided data for the review.

**Figure 1.**
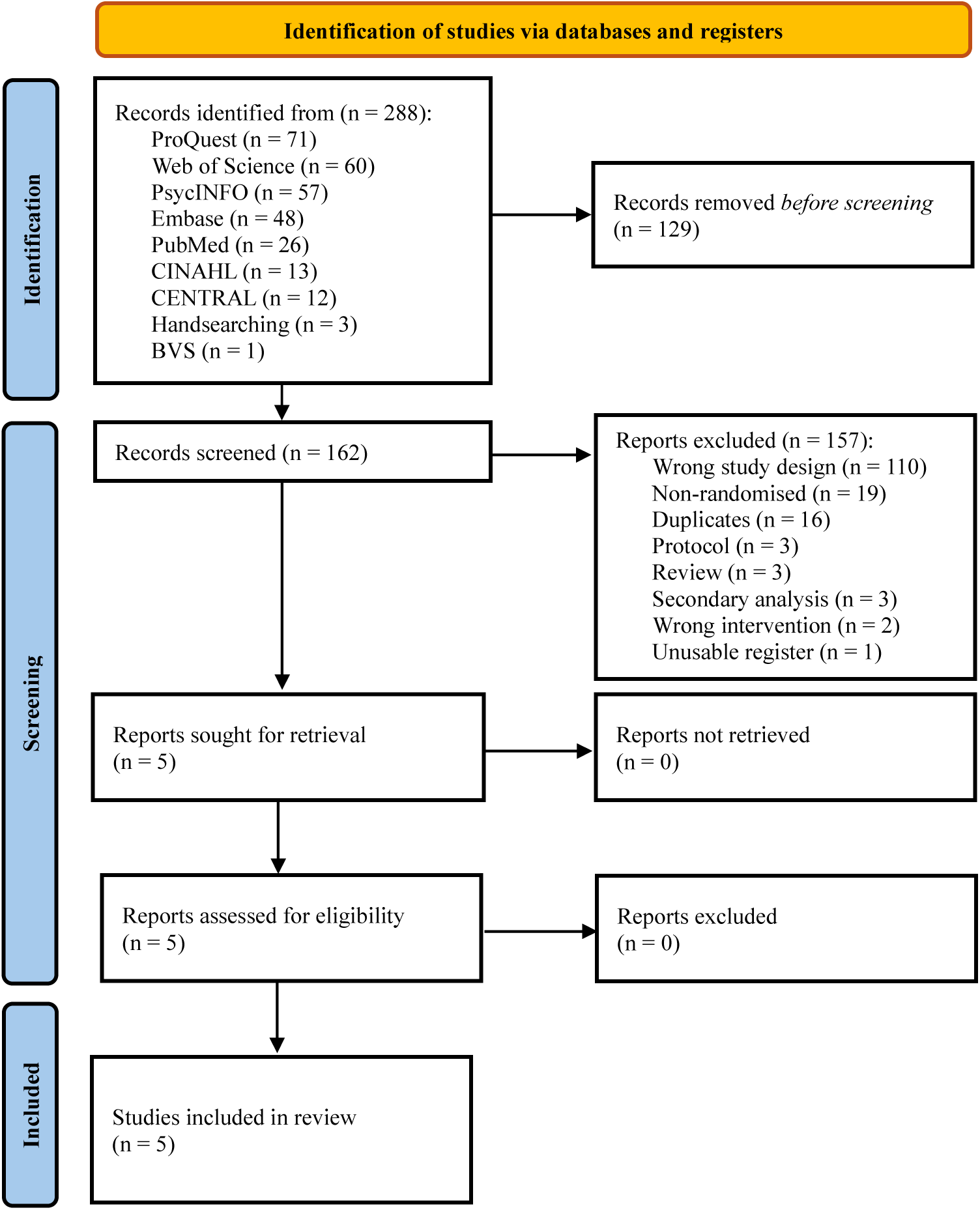
PRISMA 2020 flowchart

During the screening process, several potentially eligible articles were identified. One trial described as a RCT was excluded because allocation was based on patient preference rather than true randomisation.^17^ A clinical trial registration (ACTRN12625000718448) investigating FCT was identified but excluded due to the absence of a control group. Another registered trial (NCT05557890), aiming to test a FCT intervention in virtual reality, was also identified; however, it had not yet commenced participants recruitment. Additionally, a German study evaluating FCT within routine psychiatric clinical practice was excluded because no randomisation procedure was reported.^18^ The identification of these records reflects the breadth of the search strategy and the systematic application of predefined eligibility criteria.

Inter-reviewer agreement during title and abstract screening was high, with a proportionate agreement of 96.9%. Cohen’s kappa indicated moderate agreement between reviewers (κ = 0.53). Agreement between reviewers during full-text screening was 80% (4 agreements, 1 disagreement); Cohen’s κ could not be meaningfully interpreted due to the absence of variability in exclusion classifications, which produced degenerate marginal distributions and rendered chance-corrected agreement estimates non-informative.^19^

### 3.1. Characteristics of included studies

Details of the included trials are provided in Appendix 12, while a summary is presented in Table 1. All included trials were published in English. Most RCTs reported receiving financial support, and the mean journal impact factor across publications was 2.3 (range: not published to 3.8). All FCT trials were conducted as single-centre studies. Considering the four trials with available outcome data and excluding the duplicated sample reported across the German publications, a total of 429 participants were recruited across the RCTs, of whom 169 were randomised to an FCT-based intervention. The empty study recruited 66 people, but the record does not state how many participants are in each group; in fact, they recruited 6 more people than the planned sample size. Waitlist was the most frequently used comparator condition across trials, while one study employed a no-intervention control. The empty study identified through trial registry records planned to use group psychotherapy as the comparator condition.

**Table 1.** Summary of included studies.

| Author | Study type | Sample Size | Setting | Intervention Characteristics | Comparison | Primary Outcome | Country |
| --- | --- | --- | --- | --- | --- | --- | --- |
| 20 | Grey literature (RCT) | N = 75<br>FC = 25<br>DSD = 25<br>Control = 25 | Adults; non-clinical sample | FC group; Weekly, in-person, 1 hour per session for 6 weeks | DSD (book with structured exercises) and control (no intervention) | Time of assessment: Baseline; 8-months<br>Instrument: MFODS | England |
| 23 | RCT | N = 208<br>FCS = 104<br>WL = 104 | Adults; non-clinical sample | FCS; in person, 3-day seminar, nonrecurring | WL | Time of assessment: Baseline, 2 weeks, 4 months<br>Instrument: EXIS.pers | Germany |
| 21 | RCT | N = 80<br>FCT = 40<br>WL = 40 | Adults; non-clinical sample | FCT; Single-day workshops (pandemic-adjusted), One-time participation in a single-day workshop | WL | Time of assessment: Baseline; 1 month; 6 months<br>Instrument: BSI | Hungary |
| 22 | RCT | N = 208<br>FCS = 104<br>WL = 104 | Adults; non-clinical sample | FCS; in person, 3-day seminar, nonrecurring | WL | Time of assessment: Baseline, 2 weeks, 4 months<br>Instrument: OQ-45.2 | Germany |
| NCT06863 948 | Grey literature (Pilot RCT) | N = 66 | Adults; Subclinical, mild, or moderate anxiety/depressive symptoms | Terebenin's Method (Psychodrama-like enactments, systemic constellation exercises and body-oriented practices) | CBT, Gestalt, or another recognised approach | Time of assessment: Baseline, ~8 weeks, ~1-3 months<br>Instrument: BDI-II or HAM-D and STAI or Anxiety and HADS-A | Russia |

**Abbreviations:** RCT, randomised controlled trial; FC, Family Constellation; DSD, Do Something Different; MFODS, Multidimensional Fear of Death Scale; FCS, Family Constellation Seminars; WL, Waitlist; EXIS.pers, Experience In Social Systems Questionnaire, personal domain; FCT, Family Constellation Therapy; BSI, Brief Symptom Inventory; OQ-45.2, The Outcome Questionnaire; CBT, Cognitive-Behavioural Therapy; BDI-II, Beck Depression Inventory–II; HAM-D, Hamilton Depression Rating Scale; STAI, The State-Trait Anxiety Inventory; HADS-A, Anxiety and Depression Scale Anxiety subscale.

Goode^20^ conducted a RCT in England involving a non-clinical sample of first-semester student nurses. Participants were randomly allocated to three groups: a Do Something Different (DSD) intervention, a Family Constellations (FC) group intervention, or a no-intervention control condition (n = 25 per group). Both active interventions were delivered over a six-week period. The FC intervention consisted of a group-based FC programme, whereas the DSD group received a structured behavioural programme based on the DSD approach. The primary outcome assessed was fear of death and dying, measured using the Multidimensional Fear of Death Scale (MFODS) at baseline and at an eight-month follow-up. Adverse effects were not reported. The study was conducted in a non-clinical setting and did not report detailed inclusion/exclusion criteria or baseline demographic characteristics separately by group. No funding source was declared.

Konkolÿ Thege and Szabó^21^ conducted a RCT in Hungary to evaluate a pandemic-adjusted FCT delivered in a group setting compared with a waitlist control. Adults (≥18 years) from the general population were recruited via paid social media advertisements and were eligible if willing to attend a constellation seminar on the randomly assigned date; individuals with a current mental disorder diagnosed by a healthcare professional were excluded. Eighty participants were randomised (40 per group). The intervention consisted of a one-day, in-person group workshop (approximately 10 participants per workshop), adapted from the traditional two-to-three-day format due to COVID-19 restrictions (mask wearing and physical distancing). Two experienced providers delivered the workshops. The primary outcome was overall psychopathology measured with the Brief Symptom Inventory (BSI) at baseline, 1 month, and 6 months, and adverse effects assessed through qualitative ad hoc questions at 1 and 6 months. The study was conducted at a single centre, with funding provided by Károli Gáspár University of the Reformed Church in Hungary.

Weinhold et al.^22^ conducted a RCT in Germany evaluating a Family Constellation Seminar (FCS) compared with a waitlist control condition among a non-clinical sample of adults. Participants were randomly allocated to either the intervention or control group (n = 104 per group). The intervention consisted of a structured three-day seminar incorporating active participation and observational components, delivered in a group format. Outcomes were assessed at baseline and follow-up time points including two weeks and four months after the intervention. The primary outcome was The Outcome Questionnaire (OQ-45.2). This trial was reported across two separate journal articles: the primary report described the main outcomes of the RCT,^22^ whereas a subsequent publication presented the Experience In Social Systems Questionnaire, personal domain (EXIS.pers) as its primary outcome.^23^ Both publications were therefore considered as multiple reports of a single underlying RCT.

One additional RCT was included (NCT06863948). This pilot study evaluated an integrative group psychotherapy incorporating FC techniques for adults with subclinical to mild/moderate anxiety and depressive symptoms, using a parallel-group randomised design with an eight-week intervention format. As no outcome data were available at the time of this review, the study was included as an empty study.

### 3.2. Risk of bias

All included RCTs were judged to be at high overall risk of bias (Figure 2). The most prominent concerns arose from bias in measurement of the outcome and selection of the reported result. Despite concerns regarding the randomisation process were identified in all included RCTs, Goode’s trial allocation was based on a fixed alternating sequence;^20^ which constitutes a systematic and predictable allocation method rather than a truly random sequence. Additional concerns were observed in the trial by Konkolÿ Thege and Szabó,^21^ in which approximately half of the participants allocated to the intervention group reported prior experience with FCT compared with 21.2% in the waitlist group, suggesting a major baseline imbalance. Furthermore, three participants initially allocated to the FCT group reported illness after randomisation and were reassigned to the control group without receiving the intervention, representing a post-randomisation deviation from intended allocation. Detailed domain-level judgements and justifications are provided in Appendix 5 (Tables A4–A7). Also, a visual summary of domain-level judgements for each trial is provided in Appendix 8.

**Figure 2.**
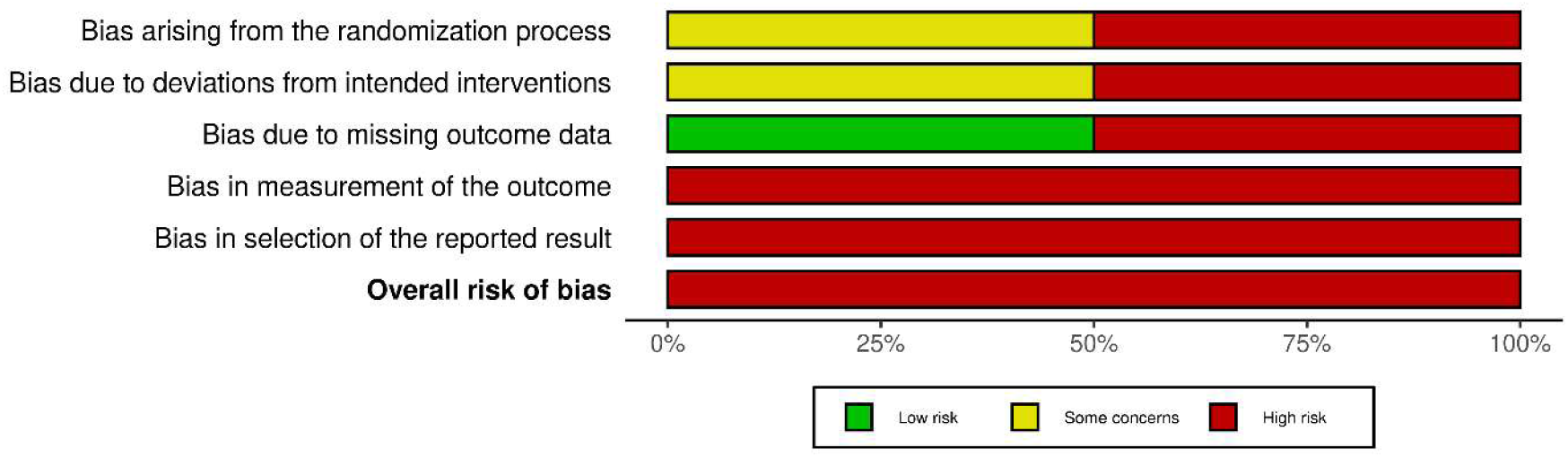
Summary of the risk of bias assessment

### 3.3. Certainty of evidence

Across all clinical comparisons, the certainty of the evidence was rated as very low. Downgrading was primarily driven by high risk of bias across included trials, substantial imprecision due to small sample sizes that did not meet the optimal information size (OIS), and inconsistency where two studies were available. Certainty ratings were judged using the pre-registered downgrading framework outlined in the publicly available protocol, ensuring independent replication of all GRADE judgements.

Overall, the body of evidence was consistently downgraded across comparisons, reflecting methodological limitations of the included trials and limited precision of the estimated effects. Detailed justifications and descriptive tables are presented in Appendix 9 (Tables A8-A11).

### 3.4. Fear of death

Fear of death was evaluated in one unpublished RCT using the MFODS at a mean follow-up of 8 months. The between-group difference was not statistically significant (MD −2.10 points; 95% CI −14.25 to 10.05; p = 0.73). This comparison was informed by a single small-sample trial (n = 17 per group in a per protocol [PP] analysis) and relied exclusively on self-reported outcomes (see Appendix 10, Analysis A1.1). Consistent with these methodological limitations, the certainty of the evidence was rated as very low.

### 3.5. Experience in social systems

Experience in social systems was evaluated in two RCTs using the EXIS.pers, including PP and an intention-to-treat analysis (ITT). A meta-analysis was conducted (Figure 3-4) because the primary outcome of one trial was reported as a secondary outcome in another included RCT.^21,23^ At postintervention follow-up (2-4 weeks), the pooled effect showed no statistically significant difference between FCS and waitlist (MD −0.16 points; 95% CI −0.80 to 0.47; p = 0.61). Similar results were observed at medium-term follow-up (4-6 months), with no significant between-group difference (MD −0.18 points; 95% CI −0.71 to 0.36; p = 0.51). These findings indicate that FCS were not more effective than waiting over time (4-6 months). Consistent with these finds, the certainty of the evidence was rated as very low.

**Figure 3.**
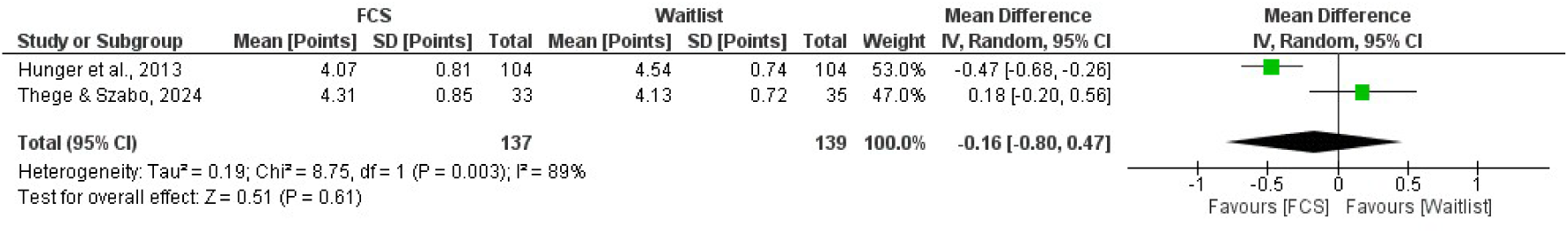
Forest plot for experience in social systems postintervention: FCS compared to WL based on a self-report scale

**Figure 4.**
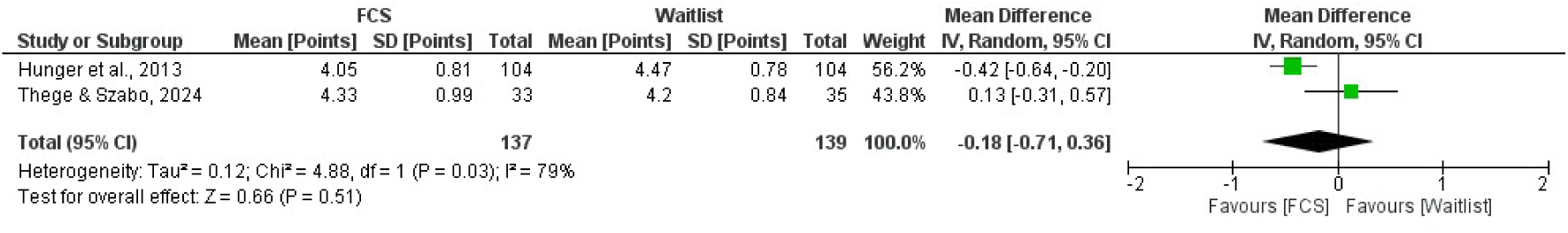
Forest plot for experience in social systems at medium-term follow-up: FCS compared to WL based on a self-report scale

For consistency with RevMan graphical conventions, effect directions were inverted (higher ir better to higher is worse) in the forest plots so that higher values consistently favoured FCT; original scale directions are preserved in the GRADE tables.

### 3.6. Overall psychopathology

Overall psychopathology was evaluated in one RCT using BSI with a PP analysis. At post-intervention follow-up, there was no statistically significant difference between FCS and waitlist (MD −0.10 points; 95% CI −0.36 to 0.16; p = 0.45). Similar findings were observed at medium-term follow-up (6 months), with no statistically significant difference between groups (MD −0.15 points; 95% CI −0.42 to 0.12; p = 0.28). These findings indicate that FCS were not more effective than waiting for reducing overall psychopathology (see Appendix 10, Analysis A2.1-2.2).

This comparison was informed by a single trial judged to be at high risk of bias and relied exclusively on self-reported outcomes. Consistent with these methodological limitations, the certainty of the evidence was rated as very low.

### 3.7. Psychological functioning

Psychological functioning was evaluated in one RCT using the OQ-45.2. At short-term follow-up (2 weeks), participants receiving FCS showed lower total OQ-45.2 scores compared with the waitlist group (MD −8.40 points; 95% CI −13.53 to −3.27; p = 0.001). A similar between-group difference was observed at medium-term follow-up (4 months), favouring FCS (MD −8.33 points; 95% CI −13.26 to −3.40; p = 0.0009).

These findings were derived from a single trial judged to be at high risk of bias and relied exclusively on self-reported outcomes. Despite the statistically significant differences observed at both time points (see Appendix 10, Analysis A3.1-3.2), the certainty of the evidence was rated as very low.

### 3.8. Adverse events

Reporting of adverse events varied substantially across studies. One trial did not describe any procedures to monitor adverse effects, and no information regarding harms was reported in either the methods or results sections.^20^ Similarly, the trial registered as NCT06863948 did not plan to collect or monitor adverse events according to the study registration, and no outcome data were available despite multiple attempts to contact the investigators.

One trial (published as two studies) used passive surveillance to monitor adverse events.^22,23^ The authors reported that no participants disclosed adverse events during the Family Constellation Seminars or throughout the 4-month study period. However, a small proportion of participants in the intervention group deteriorated over time, with deterioration observed in 3% of participants at the 2-week follow-up and 4% at the 4-month follow-up.

In contrast, Konkolÿ Thege and Szabó assessed potential iatrogenic effects using qualitative ad hoc questions.^21^ At 1-month follow-up, 36.1% of participants in the intervention group reported challenges or negative changes that they did not exclude as being related to the intervention, including worsened mood, intensified family conflicts, uncontrollable crying, incident musculoskeletal pain, and increased eating disorder symptoms. At 6-month follow-up, 25.0% of intervention participants reported possible negative changes, such as worsening family relationships, estrangement from social contacts, abdominal pain, irritability, and unwanted contact attempts by estranged family members.

In total, 139 intervention participants were included in studies that reported any information related to harms; however, reporting methods varied substantially, preventing pooled estimates.

### 3.9. Subgroup analyses

Pre-specified subgroup analyses were limited by the characteristics of the included studies. Subgroup analysis based on low risk of bias was not possible because all included trials were judged to be at high overall risk of bias. A subgroup analysis of unpublished studies was inherently represented by the fear-of-death outcome, which was informed by a single unpublished RCT.^20^ Subgroup analyses comparing ITT and PP analyses were not feasible, as only one study contributed data to each analytic.^21,23^

## 4. DISCUSSION

This systematic review found no reliable evidence to support FCT or FCT-based interventions for any clinical condition at present. Moreover, the body of evidence currently in production and expected to be published is unlikely to change the conclusions of this review, as their designs present important methodological constraints, such as pilot-study frameworks (NCT06863948) or the absence of comparison groups (ACTRN12625000718448), which limit their capacity to provide confirmatory evidence regarding clinical effectiveness.^24,25^ Other RCTs have been paused and have not started recruitment for years, according to their records (e.g., ReBEC/U1111-1284-6061; NCT05557890).

Although the pooled estimates did not demonstrate superiority of FCS over waitlist, the magnitude of the observed effects was small and centred near zero. When interpreted in a broader quantitative context, these estimates appear comparable in magnitude to the small effects typically attributed to placebo interventions. For example, the Cochrane review by Hróbjartsson and Gøtzsche reported an overall placebo effect of approximately SMD −0.23 across all clinical conditions.^26^ While direct statistical comparisons are not appropriate due to differences in outcome metrics, populations, and methodological frameworks, the point estimates observed in the present meta-analysis fall within a similar small-effect range. This contextual comparison may suggest that any observed changes associated with FCS are unlikely to exceed non-specific effects commonly observed in behavioural and placebo-controlled research. These findings are particularly important for the field of FCT, as an intervention needs to identify whether its real potential outweighs confounding biases.^27,28^ In a similar way to the classic work of Hans Eysenck, which demonstrated that psychoanalysis and other psychotherapies of his time were not superior to the mere passage of time and thereby challenged their proponents to improve methods and undertake rigorous clinical research, the present results highlight the need for methodological advancement and credible empirical evaluation within FCT-based interventions.^29^

The certainty of the evidence for all FCT comparisons assessed in RCTs was rated as very low. In addition, the small number of studies and the limited overlap of outcomes and comparators prevented the conduct of meta-analyses for more than one comparison. Together, these findings indicate that the current evidence base is insufficient not only to support clinical recommendations, but also to allow confident estimation of the magnitude or consistency of any potential effects associated with FCT-based interventions.^30^

### 4.1. Statistical power

Across the included trials, we identified issues related to statistical power and the reproducibility of sample size calculations. In several studies, the assumptions underpinning power analyses were insufficiently reported or could not be fully reproduced based just on the available information, limiting confidence in the adequacy of the planned sample sizes. These limitations may have contributed to imprecision and uncertainty in the observed effects. A detailed examination of these methodological concerns, including reproducibility checks of the reported calculations, is provided in Appendix 11. Adequately powered RCTs are essential to ensure that clinically meaningful effects can be detected with sufficient precision, reducing the risk of false findings and preventing misleading conclusions about the effectiveness or safety of an intervention.^31^

### 4.2. Comparison with prior systematic reviews

Our findings differ substantially from those reported in Konkolÿ Thege et al., who concluded that the accumulated data support the hypothesis that FCT may be an effective intervention for improving mental health in the general population.^32^ Their review included both controlled and uncontrolled studies and emphasised that nine of twelve studies reported statistically significant improvements following the intervention.^32^ In contrast, our review focused on randomised evidence and certainty of effects, which resulted in markedly more cautious conclusions.

Several methodological differences appear to account for this divergence. Notably, the random-effects meta-analysis reported by Konkolÿ Thege et al. was based on studies that varied considerably in design, population, and level of scientific scrutiny.^32^ The pooled estimate for general psychopathology incorporated data not only from controlled trials but also from quasi-experimental and uncontrolled sources, including materials that were not peer-reviewed journal articles and lacked comparison groups. Such heterogeneity in study design and evidentiary standards raises concerns regarding the interpretability of the pooled effect size and the extent to which it reflects independent, comparable clinical evidence.^33, 34^ When the evidence base is restricted to randomised comparisons and appraised using a certainty-of-evidence framework, the apparent signal of effectiveness becomes highly uncertain, with very low confidence in any estimated effects.

These differences highlight how broader inclusion of uncontrolled or methodologically heterogeneous sources may yield more optimistic quantitative estimates, whereas stricter methodological criteria prioritising internal validity and risk-of-bias considerations lead to more conservative interpretations.^34^ Rather than representing conflicting findings, the discrepancy between reviews primarily reflects divergent eligibility criteria, analytical strategies, and thresholds for evidentiary certainty.

### 4.3. Implications for the scientific community and public health decision-making

These methodological discrepancies may have broader consequences beyond academic debate, particularly for scientific communities and public policy stakeholders in contexts where FCT has been incorporated into healthcare systems, such as Brazil and Germany.^5,21^ Until now, clinical communities and policy makers may have relied on systematic reviews and evidence syntheses whose conclusions were informed by analyses with substantial methodological limitations,^32^ including the aggregation of fragile study designs and non-comparable sources that may have inflated estimated effects (discussed earlier in this review). An illustrative example is the Clinical Effectiveness of Family Constellation – Evidence Map (PAHO/BIREME), which included 16 publications and reported multiple medical and health-related outcomes while drawing largely on non-randomised designs, including uncontrolled studies, case reports, and qualitative research.^35^ Despite the limited presence of rigorous randomised evidence, this synthesis concluded that FCT may offer benefits, particularly for mental health, reinforcing its potential as a complementary therapeutic approach. Such interpretations highlight the importance of distinguishing between exploratory evidence and high-certainty clinical findings, as policy decisions grounded primarily in heterogeneous or low-certainty data may inadvertently promote premature implementation of interventions whose effectiveness and safety remain uncertain.

### 4.4. Clinical significance

Among the studies included in this review, only one RCT permitted a meaningful discussion of clinical significance, as it employed an outcome measure (OQ-45.2) with established thresholds for clinically significant change.^22,36^ Although statistically significant group-level differences were observed, the study did not demonstrate evidence of clinically meaningful individual change, with most participants failing to exceed the Reliable Change Index (RCI) threshold (14 points on OQ-45.2).^36,37^ This limitation is compounded by the fact that a substantial proportion of participants were already within the functional range at baseline, raising questions about the clinical purpose of exposing psychologically healthy individuals to a therapeutic intervention designed to treat distress or dysfunction.^38^

These considerations are particularly salient when viewed alongside issues of safety and adverse event monitoring. While the authors reported the use of passive surveillance for adverse effects,^22,23^ relying primarily on spontaneous participant reporting, such an approach may underestimate the occurrence of negative effects, especially when participants lack the conceptual framework to recognise or attribute harm to the intervention.^39,40^ Contemporary understandings of adverse effects in psychotherapy increasingly acknowledge that lack of improvement, symptom deterioration, changes in family relations or changes in the life circumstances may themselves constitute negative outcomes.^41^ In this context, exposing non-clinical participants to an intervention without clear potential for benefit raises ethical concerns grounded in the principle that, if benefit cannot be reasonably expected, harm should at minimum be avoided.^40,42,43^ Observational follow-up data also indicate evidence of deterioration at six months in a notable proportion of participants (e.g., 13.7% in wellbeing, 15.7% in presence of meaning, and 8.8% in overall psychopathology), suggesting that even exposure to a single 2-day FCT intervention may be associated with worsening in domains directly related to mental health and quality of life.^44^ Future research in FCT should benefit from instruments for monitoring and measuring adverse effects in psychotherapy.^45^

### 4.5. Strengths and limitations

This review has an important limitation that warrants consideration. The meta-analysis was conducted following a deviation from the original protocol, which had specified that only primary outcomes from RCTs would be synthesised quantitatively. However, after identifying the same outcome instrument measured across two trials, we pooled these data to estimate the potential effect of FCT. To minimise the risk of bias introduced by this deviation, all analytical steps were conducted in accordance with the pre-specified statistical framework outlined in the protocol. An additional limitation is that heterogeneity across studies was not explored in detail in the Discussion, as recommended by PRISMA 2020 (Item 20c). Despite this limitation, the review has several methodological strengths. The synthesis focused exclusively on RCTs when evaluating intervention effects, thereby prioritising internal validity and causal inference. The study protocol and statistical analysis plan were prospectively registered, and risk of bias was assessed using the RoB 2 tool, with certainty of evidence evaluated according to the GRADE framework. Reporting adhered to the PRISMA 2020 guidelines, and the full screening process, data extraction procedures, and supporting materials were transparently documented in the appendices, enhancing reproducibility and transparency.

## 5. CONCLUSION

Based on our synthesis of all available RCTs of FCT across all clinical conditions in both the published and unpublished literature, we were unable to identify reliable evidence to support the use of FCT or FCT-based interventions for any clinical indication. The current evidence base is characterised by very low certainty, substantial methodological limitations, and a lack of robust demonstrations of clinically meaningful benefit. Higher-quality RCTs, with adequate power, appropriate comparators, and rigorous monitoring of adverse effects, would be required to justify the use of FCT in real-world settings. Notably, the present review focused exclusively on clinical effectiveness and safety outcomes and did not address the methodological or theoretical limitations underlying the conceptual framework of FCT itself.^4,6^

## Data Availability

All data produced in the present work are contained in the manuscript.

## 6. ACKNOWLEDGMENTS

F.L.S. dedicates this work to Ragnar S. — rest in peace, my friend.

