## Appendices for "Family Constellations for All Clinical Conditions: A Systematic Review and Meta-analysis Showing a Lack of Supporting Evidence"

**Appendix 1** – Search strategy

**Table A1.** Search strategy, concepts and key search terms

| Search | Concept | Key search terms |
|---|---|---|
| #1 | Exposure | "Family Constellation"[Title/Abstract] OR "Family Constellations"[Title/Abstract] OR "Systemic Constellation"[Title/Abstract] OR "Systemic Constellations"[Title/Abstract] OR "System Constellation"[Title/Abstract] OR "System Constellations"[Title/Abstract] OR "Structural Constellations"[Title/Abstract] OR "Family Constellation Therapy"[Title/Abstract] OR "Systemic Constellation Therapy"[Title/Abstract] |
| #2 | Study design | randomi*ed[Title/Abstract] OR trial[Title/Abstract] OR control*[Title/Abstract] OR randomly[Title/Abstract] |
| #3 | Strategy | #1 AND #2 |

**Appendix 2** – Search strategy results

**Table A2.** Strategy search results by database

| Database searched | Date of search | Number of results |
|---|---|---|
| **ProQuest** | 27/11/2025 | 71 |
| **Web of Science** | 27/11/2025 | 60 |
| **PsycINFO via Ovid** | 27/11/2025 | 57 |
| **Embase** | 27/11/2025 | 48 |
| **PubMed** | 27/11/2025 | 26 |
| **CINAHL** | 27/11/2025 | 13 |
| **CENTRAL** | 27/11/2025 | 12 |
| **BVS** | 27/11/2025 | 1 |
| **Citation/Handsearching** | N/A | 3 |
| **Total** | | 291 |
| **After de-duplication** | | 162 |

**Note:** N/A = not applicable

**Appendix 3 –** Search strategy adaptations

**MEDILINE (PubMed)**

1) "Family Constellation"[tiab] OR "Family Constellations"[tiab] OR "Systemic Constellation"[tiab] OR "Systemic Constellations"[tiab] OR "System Constellation"[tiab] OR "System Constellations"[tiab] OR "Structural Constellations"[tiab] OR "Family Constellation Therapy"[tiab] OR "Systemic Constellation Therapy"[tiab]
2) randomi*ed[tiab] OR trial[tiab] OR control*[tiab] OR randomly[tiab]
3) #1 AND #2

**CENTRAL (Cochrane Library)**

1. "Family Constellation":ti,ab OR "Family Constellations":ti,ab OR "Systemic Constellation":ti,ab OR "Systemic Constellations":ti,ab OR "System Constellation":ti,ab OR "System Constellations":ti,ab OR "Structural Constellations":ti,ab OR "Family Constellation Therapy":ti,ab OR "Systemic Constellation Therapy":ti,ab
2. randomi*ed:ti,ab OR trial:ti,ab OR control*:ti,ab OR randomly:ti,ab
3. #1 AND #2

**PsycInfo (via Ovid)**

1. "Family Constellation".ti,ab. OR "Family Constellations".ti,ab. OR "Systemic Constellation".ti,ab. OR "Systemic Constellations".ti,ab. OR "System Constellation".ti,ab. OR "System Constellations".ti,ab. OR "Structural Constellations".ti,ab. OR "Family Constellation Therapy".ti,ab. OR "Systemic Constellation Therapy".ti,ab.
2. randomi*ed.ti,ab. OR trial.ti,ab. OR control*.ti,ab. OR randomly.ti,ab.
3. #1 AND #2

**CINHAL**

1. (TI "Family Constellation" OR AB "Family Constellation") OR (TI "Family Constellations" OR AB "Family Constellations") OR (TI "Systemic Constellation" OR AB "Systemic Constellation") OR (TI "Systemic Constellations" OR AB "Systemic Constellations") OR (TI "System Constellation" OR AB "System Constellation") OR (TI "System Constellations" OR AB "System Constellations") OR (TI "Structural Constellations" OR AB "Structural Constellations") OR (TI "Family Constellation Therapy" OR AB "Family Constellation Therapy") OR (TI "Systemic Constellation Therapy" OR AB "Systemic Constellation Therapy")
2. (TI randomi*ed OR AB randomi*ed) OR (TI trial OR AB trial) OR (TI control* OR AB control*) OR (TI randomly OR AB randomly)

3. S1 AND S2

**Embase (via Ovid)**

1. "Family Constellation".tw. OR "Family Constellations".tw. OR "Systemic Constellation".tw. OR "Systemic Constellations".tw. OR "System Constellation".tw. OR "System Constellations".tw. OR "Structural Constellations".tw. OR "Family Constellation Therapy".tw. OR "Systemic Constellation Therapy".tw.
2. randomi*ed.tw. OR trial.tw. OR control*.tw. OR randomly.tw.
3. #1 AND #2

**Web of Science**

Seach for all fields*

1. ("Family Constellation" OR "Family Constellations" OR "Systemic Constellation" OR "Systemic Constellations" OR "System Constellation" OR "System Constellations" OR "Structural Constellations" OR "Family Constellation Therapy" OR "Systemic Constellation Therapy")
2. randomi*ed OR trial OR control* OR randomly
3. #1 AND #2

**BVS databases (English)**

1. tw:"Family Constellation" OR tw:"Family Constellations" OR tw:"Systemic Constellation" OR tw:"Systemic Constellations" OR tw:"System Constellation" OR tw:"System Constellations" OR tw:"Structural Constellations" OR tw:"Family Constellation Therapy" OR tw:"Systemic Constellation Therapy"
2. tw:randomi*ed OR tw:trial OR tw:control* OR tw:randomly
3. #1 AND #2

**BVS databases (Brazilian-Portuguese)**

1. tw:"Constelação Familiar" OR tw:"Constelações Familiares" OR tw:"Constelação Sistêmica" OR tw:"Constelações Sistêmicas" OR tw:"Constelação de Sistemas" OR tw:"Constelações de Sistemas" OR tw:"Constelações Estruturais" OR tw:"Terapia de Constelação Familiar" OR tw:"Terapia de Constelação Sistêmica"
2. tw:randomizado OR tw:ensaio OR tw:controlado OR tw:aleatoriamente OR tw:aleatório OR tw:aleatorizado
3. #1 AND #2

**BVS databases (Spanish)**

1. tw:"Constelación Familiar" OR tw:"Constelaciones Familiares" OR tw:"Constelación Sistémica" OR tw:"Constelaciones Sistémicas" OR tw:"Constelación de Sistemas" OR tw:"Constelaciones de Sistemas" OR tw:"Constelaciones Estructurales" OR tw:"Terapia de Constelación Familiar" OR tw:"Terapia de Constelación Sistémica"
2. tw:aleatorizado OR tw:ensayo OR tw:controlado
3. #1 AND #2

**ProQuest**

1. TI,AB("Family Constellation") OR TI,AB("Family Constellations") OR TI,AB("Systemic Constellation") OR TI,AB("Systemic Constellations") OR TI,AB("System Constellation") OR TI,AB("System Constellations") OR TI,AB("Structural Constellations") OR TI,AB("Family Constellation Therapy") OR TI,AB("Systemic Constellation Therapy")
2. TI,AB(randomi*ed) OR TI,AB(trial) OR TI,AB(control*) OR TI,AB(randomly)
3. #1 AND #2

**International Clinical Trials Registry Platform (ICTRP)**

As the ICTRP advanced data search option does not allow more than three lines for building the search strategy, the search was performed in the standard search engine.

Since the search engine cannot generate searches with special characters (e.g. random*ed), the terms used to filter study design were written out in full.

("Family Constellation" OR "Family Constellations" OR "Systemic Constellation" OR "Systemic Constellations" OR "System Constellation" OR "System Constellations" OR "Structural Constellations" OR "Family Constellation Therapy” OR "Systemic Constellation Therapy") AND (randomized OR randomised OR trial OR control OR controlled OR controlled OR randomly)

**Appendix 4 –** PICOTS framework

**Table A3.** Strategy for comprehensive inclusion of studies.

| Domain | Criteria |
|---|---|
| Population | Any study involving humans (no restriction of age, sex, ethnicity, diagnoses, etc.) |
| Intervention | Any FCT-based intervention (e.g., FCT with horses, FCT seminars, etc.) |
| Comparison | Any control group (e.g., waitlist, gold-standard treatment, etc.) |
| Outcome | Any primary outcome |
| Timing | Any follow-up duration (e.g., post-treatment, 12 months, etc.) |
| Study design | Randomised controlled trials (e.g., pilot, parallel, crossover, cluster trials, etc.) |

**Appendix 5** – Justifications for risk of bias judgments

**Table A4.** Study 1: Goode, 2016

| **Randomisation process** | **Response** | **Justification** |
|---|---|---|
| 1.1 Was the allocation sequence random? | N | Allocation was based on a fixed alternating sequence (every third participant allocated to the same group), which constitutes a systematic and predictable allocation method rather than a truly random sequence. |
| 1.2 Was the allocation sequence concealed until participants were enrolled and assigned to interventions? | N | |
| 1.3 Did baseline differences between intervention groups suggest a problem with the randomisation process? | PY | Although demographic variables were collected at baseline (stage 1), baseline characteristics were not clearly reported by intervention group at the time of randomisation. Comparisons between groups are presented narratively and often rely on post-attrition (stage 2) data, limiting the ability to assess true baseline comparability. |
| **Deviations from the intended interventions** | **Response** | **Justification** |
| 2.1 Were participants aware of their assigned intervention during the trial? | Y | Because it was a psychotherapeutic intervention, the patients had no way of not knowing that they were receiving Constellation, DSD or nothing, and the therapists had no way of not knowing that they were treating their patients. |
| 2.2 Were carers and people delivering the interventions aware of participants' assigned intervention during the trial? | Y | |
| 2.3 [if applicable:] If Y/PY/NI to 2.1 or 2.2: Were important nonprotocol interventions balanced across interventions group? | N | Important non-protocol interventions were not balanced across groups. Participants in the intervention arms received substantially different levels of contact, activities, and exposure compared to the control group, and adherence and dose varied widely within and between intervention groups. |
| 2.4 [If applicable:] Were there failures in implementing the intervention that could have affected the outcome? | Y | There were issues in implementing the interventions, including unmeasured adherence to the DSD task and low attendance at constellation sessions, which could plausibly have affected outcomes. |
| 2.5 [If applicable:] Was there non-adherence to the assigned intervention regimen that could have affected participants' outcomes? | PY | Substantial non-adherence occurred, particularly in the constellation group, and adherence to the DSD intervention was not measured. |
| 2.6 If N/PN/NI to 2.3, or Y/PY/NI to 2.4 or 2.5: Was an appropriate analysis used to | N | No appropriate analysis was used to estimate the effect of adhering to the intervention. Adherence was not measured for the DSD intervention, substantial non-adherence occurred in |

| | | |
|---|---|---|
| estimate the effect of adhering to the intervention? | | the constellation group, and analyses did not account for exposure or adherence. |
| **Missing outcome data** | **Response** | **Justification** |
| 3.1 Were data for this outcome available for all, or nearly all, participants randomised? | N | Outcome data were available for only 55 of the 75 randomised participants. Approximately 27% of participants were missing outcome data at follow-up, which cannot be considered “nearly all” participants. |
| 3.2 If N/PN/NI to 3.1: Is there evidence that the result was not biased by missing outcome data? | N | There is no evidence that the results were not biased by missing outcome data. Analyses were restricted to participants with complete data, with no sensitivity analyses or methods to address missingness. |
| 3.3 If N/PN to 3.2: Could missingness in the outcome depend on its true value? | PY | Given the sensitive nature of the outcome and reports that the questionnaire was distressing for some participants (e.g., "Four of the students who were later interviewed stated that the questionnaires had been a shock for them"), it is plausible that missingness depended on the true value of the outcome. It is likely that participants with higher fear of death or greater distress were less likely to complete follow-up assessments, suggesting that missingness probably depended on the true outcome value. |
| 3.4 If Y/PY/NI to 3.3: Is it likely that missingness in the outcome depend on its true value? | PY | |
| **Measurement of the outcome** | **Response** | **Justification** |
| 4.1 Was the method of measuring the outcome inappropriate? | N | The outcome was measured using a validated, multidimensional self-report instrument (MFODS), applied consistently across groups and time points. The method of outcome measurement was appropriate for the construct of interest. |
| 4.2 Could measurement or ascertainment of the outcome have differed between intervention groups? | PN | There is no indication that the procedures used to measure or ascertain the outcome differed systematically between intervention groups. |
| 4.3 If N/PN/NI to 4.1 and 4.2: Were outcome assessors aware of the intervention received by study participants? | Y | Outcomes were self-reported by participants, who were aware of the intervention they received. |
| 4.4 If Y/PY/NI to 4.3: Could assessment of the outcome have been influenced by knowledge of intervention received? | PY | Given that outcomes were self-reported and participants were aware of their assigned intervention, assessment of the outcome could plausibly have been influenced by knowledge of the intervention received. Considering the subjective nature of the outcome and the reflective content of the interventions, it is likely that such awareness influenced participants’ responses, although the magnitude of this influence cannot be determined. |
| 4.5 If Y/PY/NI to 4.4: Is it likely that assessment of the outcome was influenced by knowledge of intervention received? | PY | |

| Selection of the reported result | Response | Description |
|---|---|---|
| 5.1 Were the data that produced this result analysed in accordance with a pre-specified analysis plan that was finalized before unblinded outcome data were available for analysis? | N | There was no pre-specified analysis plan finalised prior to unblinded outcome data being available. |
| 5.2 ... multiple eligible outcome measurements (e.g. scales, definitions, time points) within the outcome domain? | PN | Although the total MFODS score was extracted as the primary outcome, the original study did not pre-specify a primary outcome. The outcome domain was measured using a multidimensional scale. All outcome measurements (total score and subscales) were fully reported. |
| 5.3 ... multiple eligible analyses of the data? | PY | Multiple eligible analyses were possible, and the absence of a pre-specified analysis plan makes it likely that the reported result was selected from among several analytical options. |

| D1: High | D2: High | D3: High | D4: High | D5: High | Overall: **High** |
|---|---|---|---|---|---|

**Note:** Unpublished doctoral dissertation (grey literature).

**Table A5.** Study 2: Hunger et al., 2022

| Randomisation process | Response | Justification |
|---|---|---|
| 1.1 Was the allocation sequence random? | Y | The allocation sequence was generated using restricted randomisation with a restricted shuffled approach, as described by Schulz and Grimes (2002), and implemented by an individual not otherwise involved in the study. Although small baseline differences were observed between groups (approximately 10% for sex and 8% for marital status), these differences are compatible with chance variation and do not systematically undermine the credibility of the randomisation process. Although randomisation was carried out by an individual not otherwise involved in the study, the article does not explicitly describe procedures to conceal the allocation sequence until participants were enrolled and assigned to interventions. |
| 1.2 Was the allocation sequence concealed until participants were enrolled and assigned to interventions? | NI | |
| 1.3 Did baseline differences between intervention groups suggest a problem with the randomisation process? | N | Minor baseline differences were observed between intervention groups (e.g., approximately 10% difference in sex distribution and 8% in marital status). These differences are small, lack a consistent pattern, and are compatible with chance variation. |
| **Deviations from the intended interventions** | **Response** | **Justification** |
| 2.1 Were participants aware of their assigned intervention during the trial? | Y | Because it was a psychotherapeutic intervention, the patients had no way of not knowing that they were receiving FCS or waiting, and the therapists had no way of not knowing that they were treating their patients. |
| 2.2 Were carers and people delivering the interventions aware of participants' assigned intervention during the trial? | Y | |
| 2.3 If Y/PY/NI to 2.1 or 2.2: Were there deviations from the | Y | The article explicitly reports protocol deviations arising from the trial context, including participants attending another FCS during the study period and participants allocated to the |

| | | |
|---|---|---|
| intended intervention that arose because of the trial context? | | intervention group who did not receive the intervention as scheduled. |
| 2.4 If Y/PY to 2.3: Were these deviations likely to have affected the outcome? | PN | Although deviations from the intended intervention occurred, the study used an intention-to-treat analysis, and deviations were limited and occurred in both groups. Based on the information reported, these deviations are unlikely to have materially affected the outcome. |
| 2.5. If Y/PY/NI to 2.4: Were these deviations from intended intervention balanced between groups? | N/A | N/A |
| 2.6 Was an appropriate analysis used to estimate the effect of assignment to intervention? | Y | The study used an intention-to-treat analysis, with participants analysed according to their randomised assignment regardless of adherence or protocol deviations, which is appropriate for estimating the effect of assignment to intervention. |
| 2.7 If N/PN/NI to 2.6: Was there potential for a substantial impact (on the result) of the failure to analyse participants in the group to which they were randomized? | N/A | N/A |
| **Missing outcome data** | **Response** | **Justification** |
| 3.1 Were data for this outcome available for all, or nearly all, participants randomised? | Y | Outcome data were available for nearly all randomised participants. Loss to follow-up was limited, occurred similarly across groups, and the study used an intention-to-treat analysis, making it unlikely that missing outcome data materially affected availability of outcome data. |
| 3.2 If N/PN/NI to 3.1: Is there evidence that the result was not biased by missing outcome data? | N/A | N/A |
| 3.3 If N/PN to 3.2: Could missingness in the outcome depend on its true value? | N/A | N/A |
| 3.4 If Y/PY/NI to 3.3: Is it likely that missingness in the outcome depend on its true value? | N/A | |
| **Measurement of the outcome** | **Response** | **Justification** |
| 4.1 Was the method of measuring the outcome inappropriate? | N | The outcome was measured using a validated self-report instrument appropriate for assessing experience in social systems, applied consistently across study groups, with no indication that the measurement method itself was inappropriate. |
| 4.2 Could measurement or ascertainment of the outcome have differed between intervention groups? | N | The outcome was measured using the same self-report instrument and procedures in both intervention groups, with no indication that outcome measurement or ascertainment differed between groups. |
| 4.3 If N/PN/NI to 4.1 and 4.2: Were outcome assessors aware of the intervention received by study participants? | Y | Outcomes were self-reported by participants, who were aware of the intervention they received. |
| 4.4 If Y/PY/NI to 4.3: Could assessment of the outcome have been influenced by knowledge of intervention received? | PY | Participants in the intervention group received FCS, whereas participants in the control group were allocated to a waiting-list condition. Given the subjective nature of the outcome and its assessment by self-report, participants' awareness of their group assignment could plausibly have influenced outcome assessment. |
| 4.5 If Y/PY/NI to 4.4: Is it likely that assessment of the outcome was influenced by | PY | |

| | | |
|---|---|---|
| knowledge of intervention received? | | |
| **Selection of the reported result** | **Response** | **Description** |
| 5.1 Were the data that produced this result analysed in accordance with a pre-specified analysis plan that was finalized before unblinded outcome data were available for analysis? | N | Although the study was registered and pre-specified its objectives and outcomes, no pre-specified statistical analysis plan detailing the analytical methods was available prior to unblinded outcome data analysis. |
| 5.2 ... multiple eligible outcome measurements (e.g. scales, definitions, time points) within the outcome domain? | PN | The outcome was clearly defined, and the same outcome measurement was consistently reported for all groups, with no indication that the result was selected from among multiple eligible outcome measurements within the outcome domain. |
| 5.3 ... multiple eligible analyses of the data? | PY | Although the outcome was clearly defined, no pre-specified statistical analysis plan was reported. In the absence of a pre-specified analysis strategy, multiple eligible analyses of the data were possible. |

| | | | | | |
|---|---|---|---|---|---|
| D1: Some C. | D2: Some C. | D3: Low | D4: High | D5: High | Overall: **High** |

**Table A6.** Study 3: Thege & Szabó, 2024

| **Randomisation process** | **Response** | **Justification** |
|---|---|---|
| 1.1 Was the allocation sequence random? | Y | Although the random sequence was generated by a professional external to the team, using Research Randomizer, a web-based tool dedicated to generating random sequences using a web-based randomisation tool, the protocol does not describe any procedures to ensure allocation concealment prior to participant enrolment and assignment. |
| 1.2 Was the allocation sequence concealed until participants were enrolled and assigned to interventions? | NI | |
| 1.3 Did baseline differences between intervention groups suggest a problem with the randomisation process? | Y | A statistically significant baseline imbalance in prior experience with family/systemic constellations favoured the intervention group (45.7% vs 21.2%). Given that this intervention is rarely prescribed, prior participation reflects active interest and positive expectancy, a prognostically relevant factor, suggesting a problem with the randomisation process. |
| **Deviations from the intended interventions** | **Response** | **Justification** |
| 2.1 Were participants aware of their assigned intervention during the trial? | Y | Because it was a psychotherapeutic intervention, the patients had no way of not knowing that they were receiving constellation or waiting, and the therapists had no way of not knowing that they were treating their patients. |
| 2.2 Were carers and people delivering the interventions aware of participants' assigned intervention during the trial? | Y | |
| 2.3 [if applicable:] If Y/PY/NI to 2.1 or 2.2: Were important nonprotocol interventions balanced across interventions group? | N | Participants in the control group engaged in family/systemic constellation interventions outside the study, constituting protocol violations, while no comparable non-protocol interventions were reported in the intervention group. In addition, participants randomised to the intervention group who did not receive the intervention due to illness were reclassified into the control group post-randomisation, a |

| | | |
|---|---|---|
| | | procedure not pre-specified in the SAP, further contributing to imbalance between groups. |
| 2.4 [If applicable:] Were there failures in implementing the intervention that could have affected the outcome? | N/A | N/A |
| 2.5 [If applicable:] Was there non-adherence to the assigned intervention regimen that could have affected participants' outcomes? | Y | Participants randomised to the intervention group did not receive the intervention due to illness and were reclassified post-randomisation, while participants in the control group engaged in family/systemic constellation interventions outside the study. Given the experiential nature of the intervention, adherence is plausibly related to outcomes. |
| 2.6 If N/PN/NI to 2.3, or Y/PY/NI to 2.4 or 2.5: Was an appropriate analysis used to estimate the effect of adhering to the intervention? | N | The per-protocol analysis was based on post-randomisation exclusions and reclassification of participants according to adherence and illness, resulting in comparison of groups that were no longer defined by the original randomisation. This approach is not appropriate for estimating the effect of adhering to the intervention. |
| **Missing outcome data** | **Response** | **Justification** |
| 3.1 Were data for this outcome available for all, or nearly all, participants randomised? | N | Outcome data were not available for all, or nearly all, randomised participants due to post-randomisation exclusions, resulting in missing outcome data. |
| 3.2 If N/PN/NI to 3.1: Is there evidence that the result was not biased by missing outcome data? | N | Missing data resulted from post-randomisation exclusions related to non-adherence, illness, and protocol violations, and no analyses were reported to assess the potential impact of missing outcomes on the results. |
| 3.3 If N/PN to 3.2: Could missingness in the outcome depend on its true value? | Y | Outcome missingness could plausibly and likely depend on true outcome values, as participants were excluded or reclassified post-randomisation due to illness and non-adherence in a per-protocol analysis. |
| 3.4 If Y/PY/NI to 3.3: Is it likely that missingness in the outcome depend on its true value? | Y | |
| **Measurement of the outcome** | **Response** | **Justification** |
| 4.1 Was the method of measuring the outcome inappropriate? | N | The outcome was measured using the Brief Symptom Inventory (BSI), a validated instrument, applied uniformly across participants and time points. |
| 4.2 Could measurement or ascertainment of the outcome have differed between intervention groups? | N | Participants were compensated for completing the same set of fully online questionnaires across all study arms, with no indication that incentives or outcome ascertainment procedures differed between intervention groups. |
| 4.3 If N/PN/NI to 4.1 and 4.2: Were outcome assessors aware of the intervention received by study participants? | Y | Outcomes were self-reported by participants, who were aware of the intervention they received. |

| | | |
|---|---|---|
| 4.4 If Y/PY/NI to 4.3: Could assessment of the outcome have been influenced by knowledge of intervention received? | PY | Participants in the intervention group received systemic/family constellation, whereas participants in the control group were allocated to a waiting-list condition. Given the subjective nature of the outcome and its assessment by self-report, participants' awareness of their group assignment could plausibly have influenced outcome assessment. |
| 4.5 If Y/PY/NI to 4.4: Is it likely that assessment of the outcome was influenced by knowledge of intervention received? | PY | |
| **Selection of the reported result** | **Response** | **Description** |
| 5.1 Were the data that produced this result analysed in accordance with a pre-specified analysis plan that was finalized before unblinded outcome data were available for analysis? | Y | The analysis plan was pre-specified but described the planned analyses at a general level, without detailed specifications. |
| 5.2 ... multiple eligible outcome measurements (e.g. scales, definitions, time points) within the outcome domain? | N | The outcome was clearly defined, and the same outcome measurement was consistently reported for all groups, with no indication that the result was selected from among multiple eligible outcome measurements within the outcome domain |
| 5.3 ... multiple eligible analyses of the data? | PY | Although a pre-specified analysis plan described the general analytical approach (repeated-measures ANOVA across three time points), it did not specify a single primary contrast, prioritise time points, or predefine post hoc comparison procedures. As a result, multiple eligible analyses of the primary outcome were possible, making selective emphasis across analyses plausible. |

| | | | | | |
|---|---|---|---|---|---|
| D1: High | D2: High | D3: High | D4: High | D5: High | Overall: **High** |

**Note:** The study was conducted by a small research team, which may have limited the feasibility of implementing full separation of roles across trial procedures (e.g., intervention delivery, data collection, and data analysis), a feature commonly recommended in randomised controlled trials to reduce the risk of bias.

**Table A7.** Study 4: Weinhold et al., 2013

| **Randomisation process** | **Response** | **Justification** |
|---|---|---|
| 1.1 Was the allocation sequence random? | Y | The study used restricted randomisation implemented via a restricted shuffled approach, as described by Schulz & Grimes (2002). This method consists of generating a fixed number of allocations per group and randomly shuffling them to produce a sequence without replacement, ensuring balanced group sizes at the end of recruitment. While this represents a valid random sequence generation method, the adequacy of this approach critically depends on proper allocation concealment (e.g., use of opaque, sealed envelopes), which was not described. Therefore, allocation concealment cannot be confirmed. |
| 1.2 Was the allocation sequence concealed until participants were enrolled and assigned to interventions? | NI | |
| 1.3 Did baseline differences between intervention groups suggest a problem with the randomisation process? | N | Minor baseline differences were observed between intervention groups (e.g., approximately 10% difference in sex distribution and 8% in marital status). These differences |

| | | |
|---|---|---|
| | | are small, lack a consistent pattern, and are compatible with chance variation. |
| **Deviations from the intended interventions** | **Response** | **Justification** |
| 2.1 Were participants aware of their assigned intervention during the trial? | Y | Because it was a psychotherapeutic intervention, the patients had no way of not knowing that they were receiving FCS or waiting, and the therapists had no way of not knowing that they were treating their patients. |
| 2.2 Were carers and people delivering the interventions aware of participants' assigned intervention during the trial? | Y | |
| 2.3 If Y/PY/NI to 2.1 or 2.2: Were there deviations from the intended intervention that arose because of the trial context? | Y | The article explicitly reports protocol deviations arising from the trial context, including participants attending another FCS during the study period and participants allocated to the intervention group who did not receive the intervention as scheduled. |
| 2.4 If Y/PY to 2.3: Were these deviations likely to have affected the outcome? | PN | Although deviations from the intended intervention occurred, the study used an intention-to-treat analysis, and deviations were limited and occurred in both groups. Based on the information reported, these deviations are unlikely to have materially affected the outcome. |
| 2.5. If Y/PY/NI to 2.4: Were these deviations from intended intervention balanced between groups? | N/A | N/A |
| 2.6 Was an appropriate analysis used to estimate the effect of assignment to intervention? | Y | The study used an intention-to-treat analysis, with participants analysed according to their randomised assignment regardless of adherence or protocol deviations, which is appropriate for estimating the effect of assignment to intervention. |
| 2.7 If N/PN/NI to 2.6: Was there potential for a substantial impact (on the result) of the failure to analyse participants in the group to which they were randomized? | N/A | N/A |
| **Missing outcome data** | **Response** | **Justification** |
| 3.1 Were data for this outcome available for all, or nearly all, participants randomised? | Y | Outcome data were available for nearly all randomised participants. Loss to follow-up was limited, occurred similarly across groups, and the study used an intention-to-treat analysis, making it unlikely that missing outcome data materially affected availability of outcome data. |
| 3.2 If N/PN/NI to 3.1: Is there evidence that the result was not biased by missing outcome data? | N/A | N/A |
| 3.3 If N/PN to 3.2: Could missingness in the outcome depend on its true value? | N/A | N/A |
| 3.4 If Y/PY/NI to 3.3: Is it likely that missingness in the outcome depend on its true value? | N/A | |
| **Measurement of the outcome** | **Response** | **Justification** |
| 4.1 Was the method of measuring the outcome inappropriate? | N | The primary outcome (OQ-45) was measured using a validated instrument, applied uniformly across participants and time points. |
| 4.2 Could measurement or ascertainment of the outcome | N | The outcome was measured using the same self-report instrument and procedures in both intervention groups, with |

| | | |
|---|---|---|
| have differed between intervention groups? | | no indication that outcome measurement or ascertainment differed between groups. |
| 4.3 If N/PN/NI to 4.1 and 4.2: Were outcome assessors aware of the intervention received by study participants? | Y | Outcomes were self-reported by participants, who were aware of the intervention they received. |
| 4.4 If Y/PY/NI to 4.3: Could assessment of the outcome have been influenced by knowledge of intervention received? | PY | Participants in the intervention group received FCS, whereas participants in the control group were allocated to a waiting-list condition. Given the subjective nature of the outcome and its assessment by self-report, participants' awareness of their group assignment could plausibly have influenced outcome assessment. |
| 4.5 If Y/PY/NI to 4.4: Is it likely that assessment of the outcome was influenced by knowledge of intervention received? | PY | |
| **Selection of the reported result** | **Response** | **Description** |
| 5.1 Were the data that produced this result analysed in accordance with a pre-specified analysis plan that was finalized before unblinded outcome data were available for analysis? | N | Although the study was registered and pre-specified its objectives and outcomes, no pre-specified statistical analysis plan detailing the analytical methods was available prior to unblinded outcome data analysis. |
| 5.2 ... multiple eligible outcome measurements (e.g. scales, definitions, time points) within the outcome domain? | PN | The outcome was clearly defined, and the same outcome measurement was consistently reported for all groups, with no indication that the result was selected from among multiple eligible outcome measurements within the outcome domain. |
| 5.3 ... multiple eligible analyses of the data? | PY | Although the outcome was clearly defined, no pre-specified statistical analysis plan was reported. In the absence of a pre-specified analysis strategy, multiple eligible analyses of the data were possible. |

| | | | | | |
|---|---|---|---|---|---|
| D1: Some C. | D2: Some C. | D3: Low | D4: High | D5: High | Overall: **High** |

**Appendix 6** – Title & Abstract screening decisions

| Screening transparency spreadsheet | | | | | | |
|---|---|---|---|---|---|---|
| **ID** | **Title** | **Decision FLS** | **Reason for exclusion FLS** | **Decision NCS** | **Reason for exclusion** | **Notes** |
| 1 | The efficacy of pandemic-adjusted family/systemic constellation therapy in improving psychopathological symptoms: A randomized controlled trial. | Include | NA | Include | NA | NA |
| 2 | Efficacy and experience of system constellations in virtual reality (VR): study protocol for a randomized controlled feasibility study. | Exclude | Protocol | Exclude | Protocol | NA |
| 3 | Family constellation seminars improve psychological functioning in a general population sample: results of a randomized controlled trial. | Include | NA | Include | NA | NA |
| 4 | The experiences of living with a sibling who stutters: a preliminary study. | Exclude | Wrong study design | Exclude | Wrong study design | NA |
| 5 | Widening the lens on family processes and the development of parent-child attachment relationships. | Exclude | Wrong study design | Exclude | Wrong study design | NA |
| 6 | Childhood Sexual Abuse and Early Timing of Puberty. | Exclude | Wrong study design | Exclude | Wrong study design | NA |
| 7 | Stress, coping, and adjustment in mothers and young adolescents in single- and two-parent families. | Exclude | Wrong study design | Exclude | Wrong study design | NA |
| 8 | Effects of family constellation seminars on itch in patients with atopic dermatitis and psoriasis: A patient preference controlled trial. | Exclude | Non-randomised study | Exclude | Non-randomised study | Originally, FLS voted to include the record, but after a consensus discussion, the record was excluded. |
| 9 | Association between parental separation, childhood trauma, neuroticism, and depression: a case control study. | Exclude | Non-randomised study | Exclude | Wrong study design | NA |
| 10 | Ethical, legal, and social aspects of health technologies for home-based paediatric palliative care - a systematic review. | Exclude | Review | Exclude | Review | NA |
| 11 | Mid- and long-term effects of family constellation seminars in a general population sample: 8- and 12-month follow-up. | Exclude | Secondary analysis | Exclude | Secondary analysis | Originally, FLS voted to include the record, but after a consensus discussion, the record was excluded. |
| 12 | Improving experience in personal social systems through family constellation seminars: results of a randomized controlled trial. | Include | NA | Include | NA | NA |
| 13 | The effects of family constellation and child gender on parental use of evaluative feedback. | Exclude | Non-randomised study | Exclude | Wrong study design | NA |
| 14 | Increased health-care utilisation in international adoptees. | Exclude | Wrong study design | Exclude | Wrong study design | NA |
| 15 | Effective methods to improve recruitment and retention in school-based substance use prevention studies. | Exclude | Wrong study design | Exclude | Wrong study design | NA |
| 16 | Social class, parental education, and obesity prevalence in a study of six-year-old children in Germany. | Exclude | Wrong study design | Exclude | Wrong study design | NA |
| 17 | Relations in families with a mentally retarded child from the perspective of the siblings. | Exclude | Wrong study design | Exclude | Wrong study design | NA |

| | | | | | | |
|---|---|---|---|---|---|---|
| 18 | Human tandem-repeat-type galectins bind bacterial non-βGal polysaccharides. | Exclude | Wrong study design | Exclude | Wrong study design | NA |
| 19 | The long-term mortality impact of combined job strain and family circumstances: A life course analysis of working American mothers. | Exclude | Wrong study design | Exclude | Wrong study design | NA |
| 20 | Teen birth rates in sexually abused and neglected females. | Exclude | Wrong study design | Exclude | Wrong study design | NA |
| 21 | [Comparative analysis of sequence alignment of SH3GL1 gene as a disease candidate gene of adolescent idiopathic scoliosis]. | Exclude | Wrong study design | Exclude | Wrong study design | NA |
| 22 | Familial risk factors for adolescent suicide: a case-control study. | Exclude | Non-randomised study | Exclude | Wrong study design | NA |
| 23 | [Family constellation in mental disorder. results and methods of statistical investigations of family size, birth order and sib position (author's transl)]. | Exclude | Duplicate | Exclude | Wrong study design | NA |
| 24 | [Peculiarities of constellation of parental pairs as risk factors and resistance-factors in the formation of gastroduodenal pathology in children]. | Exclude | Wrong study design | Exclude | Wrong study design | NA |
| 25 | [The pathological changes of abdominal and peripheral arteries in familial hypercholesterolemia--the result of high-resolution color Doppler ultrasonography]. | Exclude | Wrong study design | Exclude | Wrong study design | NA |
| 26 | Monoallelic expression on autosomes may explain an unusual heritable form of pigmentary mosaicism: a historical case revisited. | Exclude | Wrong study design | Exclude | Wrong study design | NA |
| 27 | | | DUPLICATE (EXCLUDED BY COVIDENCE) | | | |
| 28 | Efficacy of System Constellations in a General Population | Exclude | Duplicate | Exclude | Duplicate | NCT01352325 |
| 29 | System Constellations in Virtual Reality (VR) | Exclude | Duplicate | Exclude | Duplicate | NCT05557890 |
| 30 | Efficacy of System Constellations in a General Population | Exclude | Duplicate | Exclude | Wrong study design | NCT01352325 |
| 31 | | | DUPLICATE (EXCLUDED BY COVIDENCE) | | | |
| 32 | | | DUPLICATE (EXCLUDED BY COVIDENCE) | | | |
| 33 | | | DUPLICATE (EXCLUDED BY COVIDENCE) | | | |
| 34 | | | DUPLICATE (EXCLUDED BY COVIDENCE) | | | |
| 35 | Family constellation and chronic pelvic pain | Exclude | Protocol | Exclude | Wrong study design | https://ensaiosclinicos.gov.br/rg/RBR-6v5cvhj |
| 36 | Familienaufstellung als Einzelintervention im Gruppensetting bei chronisch-psychosozialen Konflikten: kurz-, mittel- und langfristige Wirksamkeit Family constellations as single intervention within the group setting in chronic psychosocial conflicts: sho | Exclude | Duplicate | Exclude | Duplicate | NA |
| 37 | | | DUPLICATE (EXCLUDED BY COVIDENCE) | | | |
| 38 | | | DUPLICATE (EXCLUDED BY COVIDENCE) | | | |
| 39 | Exploring the effectiveness of individual family constellations for adult psychoemotional distress: A quasi-experimental study. | Exclude | Non-randomised study | Exclude | Wrong study design | NA |
| 40 | Insights from a sixty-four-year case of anorexia nervosa: Constancy and change in symptoms and treatment. | Exclude | Wrong study design | Exclude | Wrong study design | NA |

| | | | | | | |
|---|---|---|---|---|---|---|
| 41 | | | DUPLICATE (EXCLUDED BY COVIDENCE) | | | |
| 42 | An updated systematic review on the effectiveness of family constellation therapy. | Exclude | Review | Exclude | Review | NA |
| 43 | Adlerian theory. | Exclude | Wrong study design | Exclude | Wrong study design | NA |
| 44 | Postmodern parenthood: Psychoanalytic invariants and transformations. | Exclude | Wrong study design | Exclude | Wrong study design | NA |
| 45 | Systemic constellations applied in organisations: A systematic review. | Exclude | Review | Exclude | Review | NA |
| 46 | Family constellations as single intervention within the group setting in chronic psychosocial conflicts: short-, medium-, and long-term efficacy. | Exclude | Duplicate | Exclude | Duplicate | NA |
| 47 | | | DUPLICATE (EXCLUDED BY COVIDENCE) | | | |
| 48 | | | DUPLICATE (EXCLUDED BY COVIDENCE) | | | |
| 49 | Perks: An Original Play and Contextual Commentary. | Exclude | Wrong study design | Exclude | Wrong study design | NA |
| 50 | Parenting as mediator between post-divorce family structure and children's well-being. | Exclude | Wrong study design | Exclude | Wrong study design | NA |
| 51 | | | DUPLICATE (EXCLUDED BY COVIDENCE) | | | |
| 52 | | | DUPLICATE (EXCLUDED BY COVIDENCE) | | | |
| 53 | | | DUPLICATE (EXCLUDED BY COVIDENCE) | | | |
| 54 | | | DUPLICATE (EXCLUDED BY COVIDENCE) | | | |
| 55 | Improving adherence among adolescents with type 1 diabetes. | Exclude | Wrong study design | Exclude | Wrong study design | NA |
| 56 | Effectiveness of systemic constellations-Exploratory findings of the Heidelberg RCT study. | Exclude | Secondary analysis | Exclude | Secondary analysis | Originally, FLS voted to include the record, but after a consensus discussion, the record was excluded. |
| 57 | | | DUPLICATE (EXCLUDED BY COVIDENCE) | | | |
| 58 | | | DUPLICATE (EXCLUDED BY COVIDENCE) | | | |
| 59 | Psychological functioning in adolescent siblings of children with attention-deficit/hyperactivity disorder. | Exclude | Wrong study design | Exclude | Wrong study design | NA |
| 60 | Beyond silence and survival. | Exclude | Wrong study design | Exclude | Wrong study design | NA |
| 61 | Lay conceptions of "family": A replication and extension. | Exclude | Wrong study design | Exclude | Wrong study design | NA |
| 62 | | | DUPLICATE (EXCLUDED BY COVIDENCE) | | | |
| 63 | Family costellations in DCA: Usefulness of Family Life Space in a systemic research. | Exclude | Non-randomised study | Exclude | Wrong study design | NA |
| 64 | Adjustment of children from disrupted and adoptive homes in a residential treatment facility. | Exclude | Wrong study design | Exclude | Wrong study design | NA |
| 65 | Migraine headaches and marriage and family happiness: An ecobehavioral approach. | Exclude | Non-randomised study | Exclude | Wrong study design | NA |
| 66 | Genetic and environmental influences on maternal and sibling interaction in middle childhood: A sibling adoption study. | Exclude | Duplicate | Exclude | Wrong study design | NA |
| 67 | How dancers leave dancing: A hermeneutic investigation of the psychosocial factors in career change. | Exclude | Wrong study design | Exclude | Wrong study design | NA |
| 68 | Familial risk factors for adolescent suicide: A case-control study. | Exclude | Non-randomised study | Exclude | Wrong study design | NA |
| 69 | | | DUPLICATE (EXCLUDED BY COVIDENCE) | | | |

| | | | | | | |
|---|---|---|---|---|---|---|
| 70 | Relations of anger expression to depression and blood pressure in high school students. | Exclude | Wrong study design | Exclude | Wrong study design | NA |
| 71 | Differential parental expectations and treatment in Asian American and European American families. | Exclude | Wrong study design | Exclude | Wrong study design | NA |
| 72 | A contextual analysis of adolescent violence using the interaction model of client health behavior. | Exclude | Wrong study design | Exclude | Wrong study design | NA |
| 73 | The family correlates of maternal and paternal perceptions of differential treatment in early childhood. | Exclude | Wrong study design | Exclude | Wrong study design | NA |
| 74 | Concepts of family among special needs adoptees. | Exclude | Wrong study design | Exclude | Wrong study design | NA |
| 75 | Stressful experiences, personal and social resources, and health outcomes among married and single mothers, or, stress and the single mom. | Exclude | Wrong study design | Exclude | Wrong study design | NA |
| 76 | Behavioral and developmental characteristics of 4-6 year-old children exposed to illicit stimulant drugs in early development: A comparison of effects by nature of exposure. | Exclude | Wrong study design | Exclude | Wrong study design | NA |
| 77 | Children's emotional and behavioral functioning following the disclosure of extrafamilial sexual abuse. | Exclude | Wrong study design | Exclude | Wrong study design | NA |
| 78 | Siblings of HIV infected children: Psychosocial effects. | Exclude | Wrong study design | Exclude | Wrong study design | NA |
| 79 | The measurement of quality of life in adolescence: The Quality of Student Life Questionnaire. | Exclude | Duplicate | Exclude | Wrong study design | NA |
| 80 | Family adaptation and coping among siblings of cancer patients, their brothers and sisters, and nonclinical controls. | Exclude | Duplicate | Exclude | Wrong study design | NA |
| 81 | Problems created by the family constellation of child and adolescent psychiatric patients (from the perspective of personality characteristics). | Exclude | Non-randomised study | Exclude | Wrong study design | NA |
| 82 | Nonverbal intelligence and verbal achievement in deaf adolescents: An examination of heredity and environment. | Exclude | Wrong study design | Exclude | Wrong study design | NA |
| 83 | Social class, maternal psychopathology, and mediational processes in young children. | Exclude | Wrong study design | Exclude | Wrong study design | NA |
| 84 | Learning disorders in children: Sibling studies. | Exclude | Wrong study design | Exclude | Wrong study design | NA |
| 85 | Urban early adolescents, crowding and the neighbourhood experience: A preliminary investigation. | Exclude | Wrong study design | Exclude | Wrong study design | NA |
| 86 | Family constellations and eminence: The birth orders of Nobel Prize winners. | Exclude | Wrong study design | Exclude | Wrong study design | NA |
| 87 | Systemic approach to a temporary psychotherapeutic group of parents. | Exclude | Wrong study design | Exclude | Wrong study design | NA |
| 88 | Family constellation in mental disorder, results and methods of statistical investigations of family size, birth order and sib position. | Exclude | Wrong study design | Exclude | Wrong study design | NA |
| 89 | The effects of family size, birth order, sibling separation and crowding on the academic achievement of boys and girls. | Exclude | Wrong study design | Exclude | Wrong study design | NA |
| 90 | Familial attitudes in paranoid schizophrenics and normals from two socioeconomic classes. | Exclude | Wrong study design | Exclude | Wrong study design | NA |
| 91 | Factors influencing the growth of intelligence in young children. | Exclude | Wrong study design | Exclude | Wrong study design | NA |

| | | | | | | |
|---|---|---|---|---|---|---|
| 92 | The private practice of child psychiatry, a ten-year study. | Exclude | Non-randomised study | Exclude | Wrong study design | NA |
| 93 | Effects of verbal stimuli on autonomic responsivity of medicated and nonmedicated schizophrenics and character disorders. | Exclude | Wrong study design | Exclude | Wrong study design | NA |
| 94 | Study of Rorschach psychogrames in a training center for true mentally deficient boys and girls. | Exclude | Wrong study design | Exclude | Wrong study design | NA |
| 95 | Family constellations of "normal" and "disturbed" marriages: An empirical study. | Exclude | Non-randomised study | Exclude | Wrong study design | NA |
| 96 | | | DUPLICATE (EXCLUDED BY COVIDENCE) | | | |
| 97 | | | DUPLICATE (EXCLUDED BY COVIDENCE) | | | |
| 98 | | | DUPLICATE (EXCLUDED BY COVIDENCE) | | | |
| 99 | | | DUPLICATE (EXCLUDED BY COVIDENCE) | | | |
| 100 | | | DUPLICATE (EXCLUDED BY COVIDENCE) | | | |
| 101 | | | DUPLICATE (EXCLUDED BY COVIDENCE) | | | |
| 102 | | | DUPLICATE (EXCLUDED BY COVIDENCE) | | | |
| 103 | | | DUPLICATE (EXCLUDED BY COVIDENCE) | | | |
| 104 | | | DUPLICATE (EXCLUDED BY COVIDENCE) | | | |
| 105 | | | DUPLICATE (EXCLUDED BY COVIDENCE) | | | |
| 106 | | | DUPLICATE (EXCLUDED BY COVIDENCE) | | | |
| 107 | | | DUPLICATE (EXCLUDED BY COVIDENCE) | | | |
| 108 | | | DUPLICATE (EXCLUDED BY COVIDENCE) | | | |
| 109 | Pilot Randomized Controlled Trial of an Integrative Group Psychotherapy (Terebenin's Method) in Adults with Subclinical and Mild/Moderate Anxiety and Depression | Include | NA | Include | NA | NCT06863948 / Originally, NCS voted to exclude the record; after consensus discussion, the record was included for screening by full-text reading. |
| 110 | Efficacy and Experience of System Constellations in Virtual Reality (VR): A Randomized Control Feasibility Study | Exclude | Duplicate | Exclude | Unusable register | NCT05557890 |
| 111 | Efficacy of Family / Systemic Constellation Therapy in the General Population. A Randomized Wait-list Controlled Trial | Exclude | Duplicate | Exclude | Duplicate | NCT01352325 |
| 112 | Randomized Controlled Trial on the Efficacy of System Constellations on Psychological Well-being in a General Population Based Sample | Exclude | Duplicate | Exclude | Duplicate | NCT01352325 |
| 113 | Bench Study Assessing the Impact of the Next Generation of Vitreoretinal-Cataract System on System Set-Up and Tear-Down Time for Vitreoretinal Surgery | Exclude | Wrong study design | Exclude | Wrong study design | NA |
| 114 | | | DUPLICATE (EXCLUDED BY COVIDENCE) | | | |
| 115 | | | DUPLICATE (EXCLUDED BY COVIDENCE) | | | |
| 116 | | | DUPLICATE (EXCLUDED BY COVIDENCE) | | | |
| 117 | Different facets of addiction from a systemic perspective: a case report | Exclude | Non-randomised study | Exclude | Wrong study design | NA |
| 118 | Familial risk factors in cases of physical child maltreatment | Exclude | Wrong study design | Exclude | Wrong study design | NA |
| 119 | | | DUPLICATE (EXCLUDED BY COVIDENCE) | | | |
| 120 | | | DUPLICATE (EXCLUDED BY COVIDENCE) | | | |

| | | | | | | |
|---|---|---|---|---|---|---|
| 121 | Familienaufstellung als Einzelintervention im Gruppensetting bei chronisch-psychosozialen Konflikten: Kurz-, mittel- und langfristige Wirksamkeit Family constellations as single intervention within the group setting in chronic psychosocial conflicts: Shor | Exclude | Secondary analysis | Exclude | Secondary analysis | Originally, FLS voted to include the record, but after a consensus discussion, the record was excluded. |
| 122 | | | DUPLICATE (EXCLUDED BY COVIDENCE) | | | |
| 123 | | | DUPLICATE (EXCLUDED BY COVIDENCE) | | | |
| 124 | Beyond diagnosis: Understanding the family's role in the regulation of emotion and behavior | Exclude | Wrong study design | Exclude | Wrong study design | NA |
| 125 | | | DUPLICATE (EXCLUDED BY COVIDENCE) | | | |
| 126 | | | DUPLICATE (EXCLUDED BY COVIDENCE) | | | |
| 127 | | | DUPLICATE (EXCLUDED BY COVIDENCE) | | | |
| 128 | Human tandem-repeat-type galectins bind bacterial non-betaGal polysaccharides | Exclude | Wrong study design | Exclude | Wrong study design | NA |
| 129 | | | DUPLICATE (EXCLUDED BY COVIDENCE) | | | |
| 130 | | | DUPLICATE (EXCLUDED BY COVIDENCE) | | | |
| 131 | | | DUPLICATE (EXCLUDED BY COVIDENCE) | | | |
| 132 | | | DUPLICATE (EXCLUDED BY COVIDENCE) | | | |
| 133 | | | DUPLICATE (EXCLUDED BY COVIDENCE) | | | |
| 134 | | | DUPLICATE (EXCLUDED BY COVIDENCE) | | | |
| 135 | | | DUPLICATE (EXCLUDED BY COVIDENCE) | | | |
| 136 | | | DUPLICATE (EXCLUDED BY COVIDENCE) | | | |
| 137 | The impact of parenthood on quality of life of cancer patients | Exclude | Wrong study design | Exclude | Wrong study design | NA |
| 138 | | | DUPLICATE (EXCLUDED BY COVIDENCE) | | | |
| 139 | | | DUPLICATE (EXCLUDED BY COVIDENCE) | | | |
| 140 | | | DUPLICATE (EXCLUDED BY COVIDENCE) | | | |
| 141 | | | DUPLICATE (EXCLUDED BY COVIDENCE) | | | |
| 142 | | | DUPLICATE (EXCLUDED BY COVIDENCE) | | | |
| 143 | | | DUPLICATE (EXCLUDED BY COVIDENCE) | | | |
| 144 | | | DUPLICATE (EXCLUDED BY COVIDENCE) | | | |
| 145 | Estimation of polygenic recurrence risk: an example utilizing data for cleft lip/palate | Exclude | Wrong study design | Exclude | Wrong study design | NA |
| 146 | The role and resources of the family during the drug rehabilitation process | Exclude | Wrong study design | Exclude | Wrong study design | NA |
| 147 | Closeness to parents and the family constellation in a prospective study of five disease states: suicide, mental illness, malignant tumor, hypertension and coronary heart disease | Exclude | Wrong study design | Exclude | Wrong study design | NA |
| 148 | REACTIONS to a DISASTER | Exclude | Wrong study design | Exclude | Wrong study design | NA |
| 149 | The principal of shared responsibility of child-rearing | Exclude | Wrong study design | Exclude | Wrong study design | NA |
| 150 | | | DUPLICATE (EXCLUDED BY COVIDENCE) | | | |
| 151 | The parental attitudes of parents of child guidance cases. I. Comparisons with normals, investigations of socioeconomic and family constellation factors and relations to parents' reactions to the clinics | Exclude | Wrong study design | Exclude | Wrong study design | NA |

| | | | | | | |
|---|---|---|---|---|---|---|
| 152 | Comparison of disadvantage children with learning disabilities and their successful peer group | Exclude | Wrong study design | Exclude | Wrong study design | NA |
| 153 | The effects of physique and intrafamily tension on self-concepts in adolescent males | Exclude | Non-randomised study | Exclude | Wrong study design | NA |
| 154 | Group psychoanalysis with adult stutterers | Exclude | Wrong intervention | Exclude | Wrong study design | NA |
| 155 | The mothers of schizophrenic patients. A study of the personality and the mother-child relationship of 100 mothers and the significance of these factors in the pathogenesis of schizophrenia, in comparison with heredity | Exclude | Wrong study design | Exclude | Wrong study design | NA |
| 156 | Juvenile delinquency (comments on etiology) | Exclude | Wrong study design | Exclude | Wrong study design | NA |
| 157 | | | DUPLICATE (EXCLUDED BY COVIDENCE) | | | |
| 158 | | | DUPLICATE (EXCLUDED BY COVIDENCE) | | | |
| 159 | | | DUPLICATE (EXCLUDED BY COVIDENCE) | | | |
| 160 | | | DUPLICATE (EXCLUDED BY COVIDENCE) | | | |
| 161 | | | DUPLICATE (EXCLUDED BY COVIDENCE) | | | |
| 162 | | | DUPLICATE (EXCLUDED BY COVIDENCE) | | | |
| 163 | | | DUPLICATE (EXCLUDED BY COVIDENCE) | | | |
| 164 | | | DUPLICATE (EXCLUDED BY COVIDENCE) | | | |
| 165 | | | DUPLICATE (EXCLUDED BY COVIDENCE) | | | |
| 166 | THE EFFECTS OF FAMILY CONSTELLATION AND CHILD GENDER ON PARENTAL USE OF EVALUATIVE FEEDBACK | Exclude | Non-randomised study | Exclude | Wrong study design | NA |
| 167 | The earth outer space traffic control system constellation | Exclude | Duplicate | Exclude | Duplicate | NA |
| 168 | Earth observing system constellation design optimization through mixed integer programming | Exclude | Wrong study design | Exclude | Wrong study design | NA |
| 169 | Beyond here and there? A description and typology of multi-transnational extended family configurations of refugees | Exclude | Wrong study design | Exclude | Wrong study design | NA |
| 170 | FAMILY CONSTELLATION IN MENTAL DISORDER - RESULTS AND METHODS OF STATISTICAL INVESTIGATIONS OF FAMILY-SIZE, BIRTH-ORDER AND SIB POSITION | Exclude | Duplicate | Exclude | Wrong study design | NA |
| 171 | REDESIGNING THE FAMILY LAW SYSTEM TO PROMOTE HEALTHY FAMILIES | Exclude | Wrong study design | Exclude | Wrong study design | NA |
| 172 | THE PARENTAL ATTITUDES OF PARENTS OF CHILD-GUIDANCE CASES .1. COMPARISONS WITH NORMALS, INVESTIGATIONS OF SOCIOECONOMIC AND FAMILY CONSTELLATION FACTORS, AND RELATIONS TO PARENTS REACTIONS TO THE CLINICS | Exclude | Wrong study design | Exclude | Wrong study design | NA |
| 173 | The effect of birth order on the probability of university enrolment | Exclude | Wrong study design | Exclude | Wrong study design | NA |
| 174 | Integration as Family History? The Impact of Migration Generations on Educational Success | Exclude | Wrong study design | Exclude | Wrong study design | NA |
| 175 | | | DUPLICATE (EXCLUDED BY COVIDENCE) | | | |
| 176 | | | DUPLICATE (EXCLUDED BY COVIDENCE) | | | |
| 177 | THE MEASUREMENT OF QUALITY-OF-LIFE IN ADOLESCENCE - THE QUALITY OF STUDENT LIFE QUESTIONNAIRE | Exclude | Wrong study design | Exclude | Wrong study design | NA |

| | | | | | | |
|---|---|---|---|---|---|---|
| 178 | FAMILY ADAPTATION AND COPING AMONG SIBLINGS OF CANCER-PATIENTS, THEIR BROTHERS AND SISTERS, AND NONCLINICAL CONTROLS | Exclude | Wrong study design | Exclude | Wrong study design | NA |
| 179 | | | DUPLICATE (EXCLUDED BY COVIDENCE) | | | |
| 180 | FAMILIAL RISK-FACTORS FOR ADOLESCENT SUICIDE - A CASE-CONTROL STUDY | Exclude | Non-randomised study | Exclude | Wrong study design | NA |
| 181 | | | DUPLICATE (EXCLUDED BY COVIDENCE) | | | |
| 182 | Reflective Learning Classifier Systems for Self-Adaptive and Self-Organising Agents | Exclude | Wrong study design | Exclude | Wrong study design | NA |
| 183 | Increasing PV Penetration Level in Smartgrid System Scenario by Enhancement of Reactive Power Support using Fuzzy Logic Controller | Exclude | Wrong study design | Exclude | Wrong study design | NA |
| 184 | | | DUPLICATE (EXCLUDED BY COVIDENCE) | | | |
| 185 | | | DUPLICATE (EXCLUDED BY COVIDENCE) | | | |
| 186 | GENETIC AND ENVIRONMENTAL-INFLUENCES ON MATERNAL AND SIBLING INTERACTION IN MIDDLE CHILDHOOD - A SIBLING ADOPTION STUDY | Exclude | Wrong study design | Exclude | Wrong study design | NA |
| 187 | | | DUPLICATE (EXCLUDED BY COVIDENCE) | | | |
| 188 | Parental union dissolution and children's emotional and behavioral problems: addressing selection and considering the role of post-dissolution living arrangements† | Exclude | Wrong study design | Exclude | Wrong study design | NA |
| 189 | | | DUPLICATE (EXCLUDED BY COVIDENCE) | | | |
| 190 | | | DUPLICATE (EXCLUDED BY COVIDENCE) | | | |
| 191 | Family Structure and Children's Educational Outcomes The Role of Economic, Cultural and Social Capital | Exclude | Wrong study design | Exclude | Wrong study design | NA |
| 192 | | | DUPLICATE (EXCLUDED BY COVIDENCE) | | | |
| 193 | Satellite mapping in cities and below cities: how good is it now? | Exclude | Wrong study design | Exclude | Wrong study design | NA |
| 194 | PSYCHOLOGICAL-ASPECTS OF OBESITY IN ADOLESCENCE | Exclude | Wrong study design | Exclude | Wrong study design | NA |
| 195 | | | DUPLICATE (EXCLUDED BY COVIDENCE) | | | |
| 196 | SUOMI NPP (S-NPP) ON ORBIT PERFORMANCE SUMMARY | Exclude | Wrong study design | Exclude | Wrong study design | NA |
| 197 | Parent involvement in CBT treatment of adolescent depression: Experiences in the Treatment for Adolescents with Depression Study (TADS) | Exclude | Wrong intervention | Exclude | Wrong study design | NA |
| 198 | | | DUPLICATE (EXCLUDED BY COVIDENCE) | | | |
| 199 | | | DUPLICATE (EXCLUDED BY COVIDENCE) | | | |
| 200 | Gender, family, and healthcare during unemployment: Healthcare seeking, healthcare work, and self-sacrifice | Exclude | Wrong study design | Exclude | Wrong study design | NA |
| 201 | | | DUPLICATE (EXCLUDED BY COVIDENCE) | | | |
| 202 | Title Research on Key Technology of Testing and Verification Multi GNSS Simulator | Exclude | Wrong study design | Exclude | Wrong study design | NA |
| 203 | Next Generation CMOS Active Pixel Sensors for satellite hybrid optical communications/imaging sensor systems | Exclude | Wrong study design | Exclude | Wrong study design | NA |

| | | | | | | |
|---|---|---|---|---|---|---|
| 204 | Integration and Dimensioning of Battery Storage Systems in Commercial Building Applications with Renewable Powerplants and Battery Electric Vehicles | Exclude | Wrong study design | Exclude | Wrong study design | NA |
| 205 | | | DUPLICATE (EXCLUDED BY COVIDENCE) | | | |
| 206 | The Dynamics of Teleworking: Case Studies of Women Medical Transcriptionists from Bangalore, India | Exclude | Wrong study design | Exclude | Wrong study design | NA |
| 207 | Autonomous orbit determination for two spacecraft from relative position measurements | Exclude | Wrong study design | Exclude | Wrong study design | NA |
| 208 | | | DUPLICATE (EXCLUDED BY COVIDENCE) | | | |
| 209 | CATCH: chasing all transients constellation hunters space mission | Exclude | Duplicate | Exclude | Duplicate | NA |
| 210 | In Search of an Evidence-Based Approach to Understand and Promote Effective Parenting Practices | Exclude | Wrong study design | Exclude | Wrong study design | NA |
| 211 | Flight Performance Analysis of the CYGNSS microSatellites from On-orbit Telemetry | Exclude | Wrong study design | Exclude | Wrong study design | NA |
| 212 | | | DUPLICATE (EXCLUDED BY COVIDENCE) | | | |
| 213 | Carrier-phase differential global positioning system navigation filter for high-altitude spacecraft | Exclude | Wrong study design | Exclude | Wrong study design | NA |
| 214 | Impact of Attitude Model, Phase Wind-Up and Phase Center Variation on Precise Orbit and Clock Offset Determination of GRACE-FO and CentiSpace-1 | Exclude | Wrong study design | Exclude | Wrong study design | NA |
| 215 | Probing General Relativity and New Physics with Lunar Laser Ranging | Exclude | Wrong study design | Exclude | Wrong study design | NA |
| 216 | Creation of the new industry-standard space test of laser retroreflectors for the GNSS and LAGEOS | Exclude | Wrong study design | Exclude | Wrong study design | NA |
| 217 | Constelações Familiares no judiciário: um tema para a Psicologia? | Exclude | Wrong study design | Exclude | Review | NA |
| 218 | International Clinical Trial: What Good Can One Session Do? Exploring the Effectiveness of Family Systemic Constellation Therapy in Individual Sessions | Exclude | Non-randomised study | Exclude | Non-randomized study | Registered clinical study (ACTRN12625000718448p), without a control group. |
| 219 | | | DUPLICATE (EXCLUDED BY COVIDENCE) | | | |
| 220 | | | DUPLICATE (EXCLUDED BY COVIDENCE) | | | |
| 221 | | | DUPLICATE (EXCLUDED BY COVIDENCE) | | | |
| 222 | | | DUPLICATE (EXCLUDED BY COVIDENCE) | | | |
| 223 | | | DUPLICATE (EXCLUDED BY COVIDENCE) | | | |
| 224 | | | DUPLICATE (EXCLUDED BY COVIDENCE) | | | |
| 225 | | | DUPLICATE (EXCLUDED BY COVIDENCE) | | | |
| 226 | CATCH: Chasing All Transients Constellation Hunters Space Mission | Exclude | Wrong study design | | | NA |
| 227 | Clinical Trial: System Constellations in Virtual Reality (VR) | Exclude | Unusable register | Exclude | Unusable register | Not yet recruiting (NCT05557890) |
| 228 | Efficacy of System Constellations in a General Population | Exclude | Protocol | Exclude | Unusable register | Already published (https://doi.org/10.1111/famp.12051). Will be included. |

| | | | | | | |
|---|---|---|---|---|---|---|
| 229 | Supply Of Viscous Fluid Control (vfc) Pack For Alcon Constellation Machine. Part No 8065750957 Must Be Manufac , Soft Tip Needle 23 G Disposable , Revolution Dsp 25ga+ Ilm Forc. 6' , 27+ Ilm Forceps , Abhi System Constellation Xenon Light Source , Advance | Exclude | Wrong study design | Exclude | Wrong study design | NA |
| 230 | | | DUPLICATE (EXCLUDED BY COVIDENCE) | | | |
| 231 | | | DUPLICATE (EXCLUDED BY COVIDENCE) | | | |
| 232 | | | DUPLICATE (EXCLUDED BY COVIDENCE) | | | |
| 233 | Flight Trial Demonstration of Secure GBAS via the L-band Digital Aeronautical Communications System (LDACS) | Exclude | Wrong study design | Exclude | Wrong study design | NA |
| 234 | | | DUPLICATE (EXCLUDED BY COVIDENCE) | | | |
| 235 | | | DUPLICATE (EXCLUDED BY COVIDENCE) | | | |
| 236 | | | DUPLICATE (EXCLUDED BY COVIDENCE) | | | |
| 237 | | | DUPLICATE (EXCLUDED BY COVIDENCE) | | | |
| 238 | | | DUPLICATE (EXCLUDED BY COVIDENCE) | | | |
| 239 | | | DUPLICATE (EXCLUDED BY COVIDENCE) | | | |
| 240 | Satellite mapping in cities and below cities: how good is it now? | Exclude | Wrong study design | | | NA |
| 241 | | | DUPLICATE (EXCLUDED BY COVIDENCE) | | | |
| 242 | Systemic Constellations in Diversity Management | Exclude | Non-randomised study | Exclude | Wrong study design | NA |
| 243 | | | DUPLICATE (EXCLUDED BY COVIDENCE) | | | |
| 244 | | | DUPLICATE (EXCLUDED BY COVIDENCE) | | | |
| 245 | | | DUPLICATE (EXCLUDED BY COVIDENCE) | | | |
| 246 | | | DUPLICATE (EXCLUDED BY COVIDENCE) | | | |
| 247 | Obscured Asymmetric Crypto-Functions for Secured Identification | Exclude | Wrong study design | Exclude | Wrong study design | NA |
| 248 | | | DUPLICATE (EXCLUDED BY COVIDENCE) | | | |
| 249 | | | DUPLICATE (EXCLUDED BY COVIDENCE) | | | |
| 250 | | | DUPLICATE (EXCLUDED BY COVIDENCE) | | | |
| 251 | | | DUPLICATE (EXCLUDED BY COVIDENCE) | | | |
| 252 | Emerging advances in small-gauge retina surgery maximize outcomes | Exclude | Wrong study design | Exclude | Wrong study design | NA |
| 253 | | | DUPLICATE (EXCLUDED BY COVIDENCE) | | | |
| 254 | | | DUPLICATE (EXCLUDED BY COVIDENCE) | | | |
| 255 | | | DUPLICATE (EXCLUDED BY COVIDENCE) | | | |
| 256 | USAF KICKS OFF AFSCN EQUIPMENT MOVE FROM COLORADO TO GREENLAND | Exclude | Wrong study design | Exclude | Wrong study design | NA |
| 257 | | | DUPLICATE (EXCLUDED BY COVIDENCE) | | | |
| 258 | | | DUPLICATE (EXCLUDED BY COVIDENCE) | | | |
| 259 | | | DUPLICATE (EXCLUDED BY COVIDENCE) | | | |
| 260 | | | DUPLICATE (EXCLUDED BY COVIDENCE) | | | |
| 261 | | | DUPLICATE (EXCLUDED BY COVIDENCE) | | | |
| 262 | | | DUPLICATE (EXCLUDED BY COVIDENCE) | | | |
| 263 | | | DUPLICATE (EXCLUDED BY COVIDENCE) | | | |
| 264 | Global Positioning System Constellation Clock Performance | Exclude | Wrong study design | Exclude | Wrong study design | NA |

| | | | | | | |
|---|---|---|---|---|---|---|
| 265 | GLOBAL POSITIONING SYSTEM CONSTELLATION CLOCK PERFORMANCE | Exclude | Duplicate | Exclude | Wrong study design | NA |
| 266 | The Eearth outer space traffic control system constellation | Exclude | Wrong study design | Exclude | Wrong study design | NA |
| 267 | What will commercial satellite communications do for the military after next?. | Exclude | Wrong study design | Exclude | Wrong study design | NA |
| 268 | | | DUPLICATE (EXCLUDED BY COVIDENCE) | | | |
| 269 | | | DUPLICATE (EXCLUDED BY COVIDENCE) | | | |
| 270 | The new space race | Exclude | Wrong study design | Exclude | Wrong study design | NA |
| 271 | | | DUPLICATE (EXCLUDED BY COVIDENCE) | | | |
| 272 | | | DUPLICATE (EXCLUDED BY COVIDENCE) | | | |
| 273 | | | DUPLICATE (EXCLUDED BY COVIDENCE) | | | |
| 274 | TAOS (S80): A (LEO-MSS) system for position reporting and telemanagement services (TAOS (S80): Un systeme LEO-MSS pour des services de localisation et de telegestion) | Exclude | Wrong study design | Exclude | Wrong study design | NA |
| 275 | A small satellite constellation for many uses | Exclude | Wrong study design | Exclude | Wrong study design | NA |
| 276 | | | DUPLICATE (EXCLUDED BY COVIDENCE) | | | |
| 277 | | | DUPLICATE (EXCLUDED BY COVIDENCE) | | | |
| 278 | Vietnamese Amerasians: Practical Implications of Current Research | Exclude | Wrong study design | Exclude | Wrong study design | NA |
| 279 | Single Mothers by Choice and Inwedlock Mothers: Sex-Role Orientation, Locus of Control, and Social Support | Exclude | Wrong study design | Exclude | Wrong study design | NA |
| 280 | Stress and Psychological Symptoms in Single and Dual Parent Families | Exclude | Wrong study design | Exclude | Wrong study design | NA |
| 281 | | | DUPLICATE (EXCLUDED BY COVIDENCE) | | | |
| 282 | CAN CHILD FATALITIES, END PRODUCT OF CHILD ABUSE, BE PREVENTED? | Exclude | Wrong study design | Exclude | Wrong study design | NA |
| 283 | The Father's Impact on Mother and Child | Exclude | Wrong study design | Exclude | Wrong study design | NA |
| 284 | | | DUPLICATE (EXCLUDED BY COVIDENCE) | | | |
| 285 | | | DUPLICATE (EXCLUDED BY COVIDENCE) | | | |
| 286 | | | DUPLICATE (EXCLUDED BY COVIDENCE) | | | |
| 287 | DISCRIMINATING ADOLESCENT MALE DELINQUENTS THROUGH THE USE OF KINETIC FAMILY DRAWINGS | Exclude | Wrong study design | Exclude | Wrong study design | NA |
| 288 | UNIVERSITY-POLICE COOPERATIVE APPROACH TO JUVENILE DIVERSION - EVALUATING ITS APPLICABILITY AND EFFECTIVENESS | Exclude | Wrong study design | Exclude | Wrong study design | NA |
| Handsearch | Heilt Demut, wo Schicksal wirkt? Evaluationsstudie zu Effekten des Familien-Stellens nach Bert Hellinger | Exclude | Non-randomised study | Exclude | Wrong study design | Further confirmation that this work is not an RCT: https://doi.org/10.1177/1066480719868706 |
| Handsearch | Enhancing the Affective Domain in Order to Reduce Fear of Death in First-Year Student Nurses | Include | NA | Include | NA | NA |
| Handsearch | Familienaufstellungen in der Psychiatrischen Tagesklinik | Exclude | Non-randomised study | Exclude | Non-randomized study | NA |

**Appendix 7** – Full text screening decisions

| Screening transparency spreadsheet | | | | |
|---|---|---|---|---|
| **ID** | **Title** | **Decision** | **Reason for exclusion** | **Notes** |
| 109 | Pilot Randomized Controlled Trial of an Integrative Group Psychotherapy (Terebenin's Method) in Adults with Subclinical and Mild/Moderate Anxiety and Depression | Include | NA | Included (NCT06863948). Results were requested from the principal investigator of the registered trial (January 12, 2026). With no response, data was requested again (January 19, 2026). With no response, data was requested again (February 5, 2026). If the data is not shared, the study will remain included as an empty study. |
| 12 | Improving experience in personal social systems through family constellation seminars: results of a randomized controlled trial. | Include | NA | NA |
| 1 | The efficacy of pandemic-adjusted family/systemic constellation therapy in improving psychopathological symptoms: A randomized controlled trial. | Include | NA | NA |
| 3 | Family constellation seminars improve psychological functioning in a general population sample: results of a randomized controlled trial. | Include | NA | NA |
| Handsearch | Enhancing the Affective Domain in Order to Reduce Fear of Death in First-Year Student Nurses | Include | NA | NA |

**Appendix 8** – Risk of bias assessment of randomised evidence in FCT.

**Figure A1.** Individual risk of bias assessments of the studies included in the review.

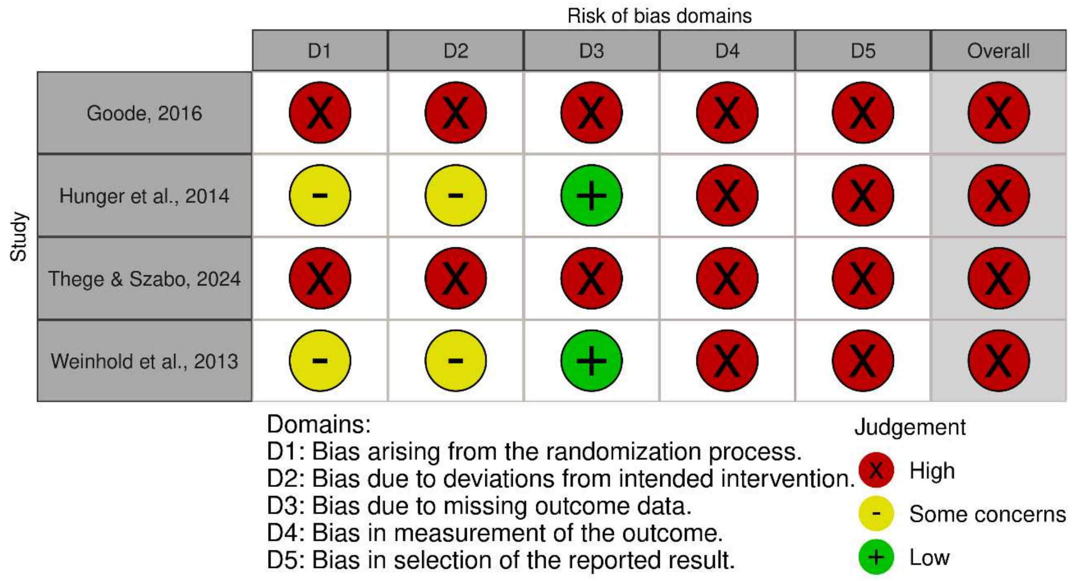

**Appendix 9** – Certainty of the evidence for the comparisons included in the review.

**Table A8.** Should FCT compared to control be used for Fear of death?

| Certainty assessment | | | | | | | № of patients | | Effect | | Certainty | Importance |
|---|---|---|---|---|---|---|---|---|---|---|---|---|
| № of studies | Study design | Risk of bias | Inconsistency | Indirectness | Imprecision | Other considerations | [Family Constellation Therapy] | [Control] | Relative (95% CI) | Absolute (95% CI) | | |
| **Fear of death (follow-up: mean 8 months; assessed with: Multidimensional Fear of Death Scale (MFODS; 42-item, 5-point Likert scale; range 42–210))** | | | | | | | | | | | | |
| 1 | randomised trials | very serious[a] | serious[b] | not serious | very serious[c] | none | 17 | 17 | - | MD **2.1 points lower** (14.25 lower to 10.05 higher) | ⨁◯◯◯ Very low[a,b,c] | IMPORTANT |

**CI:** confidence interval; **MD:** mean difference

### *Explanations*

a. Risk of bias was rated as very serious because the body of evidence was informed by a single study judged to be at high risk of bias across all domains of the RoB 2 tool.

b. Inconsistency was rated as serious because the body of evidence consisted of a single study, precluding assessment of variability in effect estimates across studies and preventing evaluation of consistency.

c. Imprecision was rated as very serious because the body of evidence consisted of a single study that did not meet the optimal information size, with fewer than 50 participants per arm. Small sample sizes are associated with increased random error and a higher likelihood of exaggerated or spurious effects.

**Table A9.** Should FCS compared to waitlist be used for Experience in social systems?

| Certainty assessment | | | | | | | № of patients | | Effect | | Certainty | Importance |
|---|---|---|---|---|---|---|---|---|---|---|---|---|
| № of studies | Study design | Risk of bias | Inconsistency | Indirectness | Imprecision | Other considerations | [Family Constellation Seminars] | [Waitlist] | Relative (95% CI) | Absolute (95% CI) | | |
| **Experience in social systems (follow-up: range 2 weeks to 4 weeks; assessed with: Experience in Social Systems Questionnaire – personal domain (EXIS.pers; 12 items, 6-point Likert scale; mean score))** | | | | | | | | | | | | |
| 2 | randomised trials | serious[a] | serious[b] | not serious | serious[c] | none | 139 | 137 | - | MD **0.16 points higher** (0.47 lower to 0.8 higher) | ⨁◯◯◯ Very low[a,b,c] | IMPORTANT |
| **Experience in social systems (follow-up: range 4 months to 6 months; assessed with: Experience in Social Systems Questionnaire – personal domain (EXIS.pers; 12 items, 6-point Likert scale; mean score))** | | | | | | | | | | | | |
| 2 | randomised trials | serious[d] | serious[e] | not serious | serious[f] | none | 139 | 137 | - | MD **0.18 points higher** (0.36 lower to 0.71 higher) | ⨁◯◯◯ Very low[d,e,f] | IMPORTANT |

**CI:** confidence interval; **MD:** mean difference

### *Explanations*

a. Risk of bias was rated as serious because one included study was judged to be at high risk of bias across all RoB 2 domains, while the other study was judged to be at high risk of bias overall, with concerns related to the randomisation process and deviations from intended interventions. These methodological limitations reduce confidence in the estimated effect.

b. Inconsistency was rated as serious due to substantial heterogeneity between studies, with markedly different point estimates in opposite directions and a high level of statistical heterogeneity ($I^2$ = 89%).

c. Imprecision was rated as serious because the total sample size did not meet the optimal information size (OIS), as recommended for continuous outcomes.

d. Risk of bias was rated as serious because one included study was judged to be at high risk of bias across all RoB 2 domains, while the other study was judged to be at high risk of bias overall, with concerns related to the randomisation process and deviations from intended interventions. These methodological limitations reduce confidence in the estimated effect.

e. Inconsistency was rated as serious due to substantial heterogeneity between studies, with markedly different point estimates in opposite directions and a high level of statistical heterogeneity ($I^2$ = 79%).

f. Imprecision was rated as serious because the total sample size did not meet the OIS, as recommended for continuous outcomes.

**Table A10.** Should FCS compared to waitlist be used for Overall psychopathology?

| Certainty assessment | | | | | | | № of patients | | Effect | | Certainty | Importance |
|---|---|---|---|---|---|---|---|---|---|---|---|---|
| № of studies | Study design | Risk of bias | Inconsistency | Indirectness | Imprecision | Other considerations | [Family Constellation Seminars] | [Waitlist] | Relative (95% CI) | Absolute (95% CI) | | |
| **Overall psychopathology (follow-up: mean 4 weeks; assessed with: Brief Symptom Inventory (BSI; Global Severity Index, 53 items, 5-point Likert scale; mean score, range 0–4))** | | | | | | | | | | | | |
| 1 | randomised trials | very serious[a] | serious[b] | not serious | very serious[c] | none | 35 | 33 | - | MD **0.1 points lower** (0.36 lower to 0.16 higher) | ⊕◯◯◯ Very low[a,b,c] | CRITICAL |
| **Overall psychopathology (follow-up: mean 6 months; assessed with: Brief Symptom Inventory (BSI; Global Severity Index, 53 items, 5-point Likert scale; mean score, range 0–4))** | | | | | | | | | | | | |
| 1 | randomised trials | very serious[d] | serious[e] | not serious | very serious[f] | none | 35 | 33 | - | MD **0.15 points lower** (0.42 lower to 0.12 higher) | ⊕◯◯◯ Very low[d,e,f] | CRITICAL |

**CI:** confidence interval; **MD:** mean difference

### *Explanations*

a. Risk of bias was rated as very serious because the body of evidence was informed by a single study judged to be at high risk of bias across all domains of the RoB 2 tool.

b. Inconsistency was rated as serious because the body of evidence consisted of a single study, precluding assessment of variability in effect estimates across studies and preventing evaluation of consistency.

c. Imprecision was rated as very serious because the body of evidence consisted of a single study that did not meet the optimal information size, with fewer than 50 participants per arm. Small sample sizes are associated with increased random error and a higher likelihood of exaggerated or spurious effects.

d. Risk of bias was rated as very serious because the body of evidence was informed by a single study judged to be at high risk of bias across all domains of the RoB 2 tool.

e. Inconsistency was rated as serious because the body of evidence consisted of a single study, precluding assessment of variability in effect estimates across studies and preventing evaluation of consistency.

f. Imprecision was rated as very serious because the body of evidence consisted of a single study that did not meet the optimal information size, with fewer than 50 participants per arm. Small sample sizes are associated with increased random error and a higher likelihood of exaggerated or spurious effects.

**Table A11.** Should FCS compared to waitlist be used for Psychological functioning?

| Certainty assessment | | | | | | | № of patients | | Effect | | Certainty | Importance |
|---|---|---|---|---|---|---|---|---|---|---|---|---|
| № of studies | Study design | Risk of bias | Inconsistency | Indirectness | Imprecision | Other considerations | [Family Constellation Seminars] | [Waitlist] | Relative (95% CI) | Absolute (95% CI) | | |
| **Psychological functioning (follow-up: mean 2 weeks; assessed with: Outcome Questionnaire-45.2 (OQ-45.2; 45 items, 5-point Likert scale; total score, range 0–180))** | | | | | | | | | | | | |
| 1 | randomised trials | serious[a] | serious[b] | not serious | serious[c] | none | 104 | 104 | - | MD **8.4 points lower** (13.53 lower to 3.27 lower) | ⨁◯◯◯ Very low[a,b,c] | CRITICAL |
| **Psychological functioning (follow-up: mean 4 months; assessed with: Outcome Questionnaire-45.2 (OQ-45.2; 45 items, 5-point Likert scale; total score, range 0–180))** | | | | | | | | | | | | |
| 1 | randomised trials | serious[d] | serious[e] | not serious | serious[f] | none | 104 | 104 | - | MD **8.33 points lower** (13.26 lower to 3.4 lower) | ⨁◯◯◯ Very low[d,e,f] | CRITICAL |

**CI:** confidence interval; **MD:** mean difference

### *Explanations*

a. Risk of bias was rated as serious because this comparison is based on a single study that was judged to be at high risk of bias overall, with major concerns related to the randomisation process and deviations from the intended intervention.

b. Inconsistency was rated as serious because the body of evidence consisted of a single study, precluding assessment of variability in effect estimates across studies and preventing evaluation of consistency.

c. Imprecision was rated as serious because the evidence was derived from a single study and did not meet the optimal information size (OIS), as recommended for continuous outcomes.

d. Risk of bias was rated as serious because this comparison is based on a single study that was judged to be at high risk of bias overall, with major concerns related to the randomisation process and deviations from the intended intervention.

e. Inconsistency was rated as serious because the body of evidence consisted of a single study, precluding assessment of variability in effect estimates across studies and preventing evaluation of consistency.

f. Imprecision was rated as serious because the evidence was derived from a single study and did not meet the OIS, as recommended for continuous outcomes.

**Appendix 10** – Data analyses for single study comparisons

**Analysis A1.1.** Fear of death (8-months)

| | FCT | | | Control | | | | Mean Difference |
|---|---|---|---|---|---|---|---|---|
| Study or Subgroup | Mean [Points] | SD [Points] | Total | Mean [Points] | SD [Points] | Total | Weight | IV, Fixed, 95% CI |
| Goode, 2016 | 109.9 | 15.88 | 17 | 112 | 20.04 | 17 | 100.0% | -2.10 [-14.25, 10.05] |
| **Total (95% CI)** | | | **17** | | | **17** | **100.0%** | **-2.10 [-14.25, 10.05]** |

Heterogeneity: Not applicable
Test for overall effect: Z = 0.34 (P = 0.73)

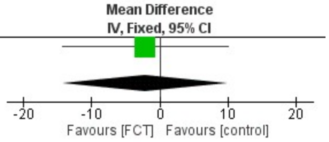

**Analysis A2.1.** Overall psychopathology (4-weeks)

| | FCS | | | Waitlist | | | | Mean Difference |
|---|---|---|---|---|---|---|---|---|
| Study or Subgroup | Mean [Points] | SD [Points] | Total | Mean [Points] | SD [Points] | Total | Weight | IV, Fixed, 95% CI |
| Thege & Szabo, 2024 | 0.61 | 0.36 | 35 | 0.71 | 0.68 | 33 | 100.0% | -0.10 [-0.36, 0.16] |
| **Total (95% CI)** | | | **35** | | | **33** | **100.0%** | **-0.10 [-0.36, 0.16]** |

Heterogeneity: Not applicable
Test for overall effect: Z = 0.75 (P = 0.45)

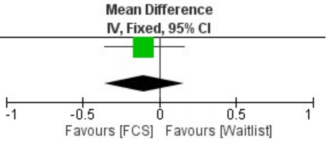

**Analysis A2.2.** Overall psychopathology (6-months)

| | FCS | | | Waitlist | | | | Mean Difference |
|---|---|---|---|---|---|---|---|---|
| Study or Subgroup | Mean [Points] | SD [Points] | Total | Mean [Points] | SD [Points] | Total | Weight | IV, Fixed, 95% CI |
| Thege & Szabo, 2024 | 0.65 | 0.4 | 35 | 0.8 | 0.7 | 33 | 100.0% | -0.15 [-0.42, 0.12] |
| **Total (95% CI)** | | | **35** | | | **33** | **100.0%** | **-0.15 [-0.42, 0.12]** |

Heterogeneity: Not applicable
Test for overall effect: Z = 1.08 (P = 0.28)

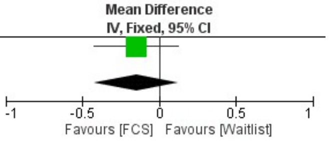

**Analysis A3.1.** Psychological functioning (2-weeks)

| | FCS | | | Waitlist | | | | Mean Difference |
|---|---|---|---|---|---|---|---|---|
| Study or Subgroup | Mean [Points] | SD [Points] | Total | Mean [Points] | SD [Points] | Total | Weight | IV, Fixed, 95% CI |
| Weinhold et al., 2013 | 42.63 | 17.39 | 104 | 51.03 | 20.22 | 104 | 100.0% | -8.40 [-13.53, -3.27] |
| **Total (95% CI)** | | | **104** | | | **104** | **100.0%** | **-8.40 [-13.53, -3.27]** |

Heterogeneity: Not applicable
Test for overall effect: Z = 3.21 (P = 0.001)

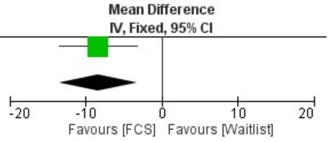

**Analysis A3.2.** Psychological functioning (4-months)

| | FCS | | | Waitlist | | | | Mean Difference |
|---|---|---|---|---|---|---|---|---|
| Study or Subgroup | Mean [Points] | SD [Points] | Total | Mean [Points] | SD [Points] | Total | Weight | IV, Fixed, 95% CI |
| Weinhold et al., 2013 | 42.46 | 17.4 | 104 | 50.79 | 18.83 | 104 | 100.0% | -8.33 [-13.26, -3.40] |
| **Total (95% CI)** | | | **104** | | | **104** | **100.0%** | **-8.33 [-13.26, -3.40]** |

Heterogeneity: Not applicable
Test for overall effect: Z = 3.31 (P = 0.0009)

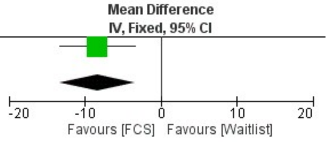

**Appendix 11** – Sample size and statistical power issues

Below we argue that all RCTs on FCT have issues related to sample size calculation and statistical power that need to be observed.

Study 1 – Goode (2016)

The doctoral dissertation by Goode (2016) did not report an a priori sample size calculation or statistical power justification. Notably, the author explicitly acknowledged that a substantially larger sample would have been necessary to achieve greater validity and reliability of the findings (Goode, 2016, p. 155).

The lack of a predefined sample size rationale, combined with acknowledged limitations in sample magnitude and data loss, suggests that the study may have been underpowered to reliably detect intervention effects.

Study 2 – Hunger et al. (2014)

Hunger and colleague's RCT did not report any a priori sample size calculation or statistical power analysis. However, because this study represents the same trial described in Weinhold et al. (2013), considerations regarding statistical power and sample size will be addressed jointly in the evaluation of that trial.

Study 3 – Konkolÿ Thege & Szabó (2024)

An independent replication of the a priori power analysis reported by Thege & Szabó (2024) was conducted using G*Power 3.1.9.7. The aim was to determine whether the reported required sample size (N = 78; 39 participants per arm) could be reproduced based solely on the information provided in the manuscript. The original article stated that the "G-Power software" was used, performing a "repeated measures mixed-design ANOVA", targeting a medium effect size (cf. Hedges' g = 0.531), with 80% power and α = 0.05.

Because the manuscript did not provide sufficient details to directly reproduce the calculation, several plausible statistical configurations were tested iteratively in G*Power.

1. Repeated measures within–between interaction

The first attempt followed the literal description of a mixed-design repeated measures ANOVA by selecting:

- F tests → ANOVA: Repeated measures, within–between interaction
- A priori: Compute required sample size – given α, power, and effect size
- Effect size f = 0.25 (medium)

- α = 0.05
- Power = 0.80
- Two groups
- Three measurement occasions (baseline, 4-week, 6-month follow-up)
- Correlation among repeated measures = 0.5
- Nonsphericity correction ε = 1

**Figure A2.** Study 3 - central and noncentral distributions, first attempt

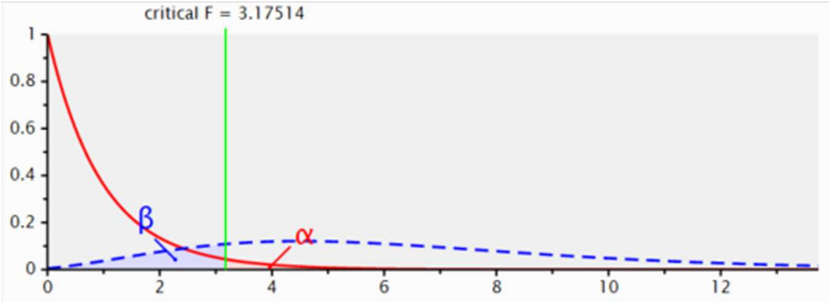

This configuration produced a smaller required sample size (N ≈ 28), indicating that the reported N = 78 was unlikely to have been based on the interaction effect.

2. Independent samples t-test using the reported Hedges' g

Given the explicit reference to Hedges' g = 0.531, a second attempt used:

- t tests → Means: Difference between two independent means (two groups)
- A priori: Compute required sample size – given α, power, and effect size
- Effect size d ≈ 0.531
- Two-tailed
- α = 0.05
- Power = 0.80
- Allocation ratio N2/N1 = 1

**Figure A3.** Study 3 - central and noncentral distributions, second attempt

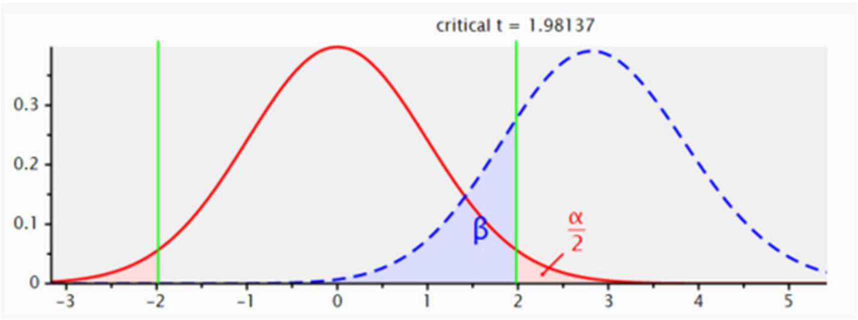

This approach yielded a much larger required sample (N ≈ 114), again inconsistent with the reported N = 78.

3. Fixed-effects ANOVA models

Additional attempts were conducted using:

- F tests → ANOVA: Fixed effects, special, main effects and interactions
- A priori: Compute required sample size – given α, power, and effect size
- Effect size f = 0.25 (medium)
- α = 0.05
- Power = 0.80
- Numerator df = 1
- Two groups

**Figure A4.** Study 3 - central and noncentral distributions, third attempt

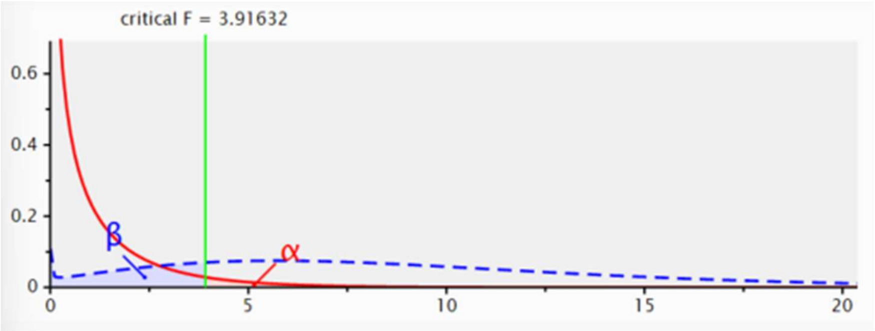

These models produced required sample sizes ranging from approximately N = 128 to N = 158, depending on numerator degrees of freedom (1 and 2, respectively), further indicating that this was not the underlying calculation.

4. Repeated measures ANOVA — between-factors effect

After iterative testing guided by the structure of the reported design, the calculation that reproduced the published sample size was:

- F tests → ANOVA: Repeated measures, between factors
- A priori: Compute required sample size – given α, power, and effect size
- Effect size f = 0.25
- α = 0.05
- Power = 0.80
- Two groups
- Number of measurements = 3
- Correlation among repeated measures r = 0.40

**Figure A5.** Study 3 - central and noncentral distributions, fourth attempt

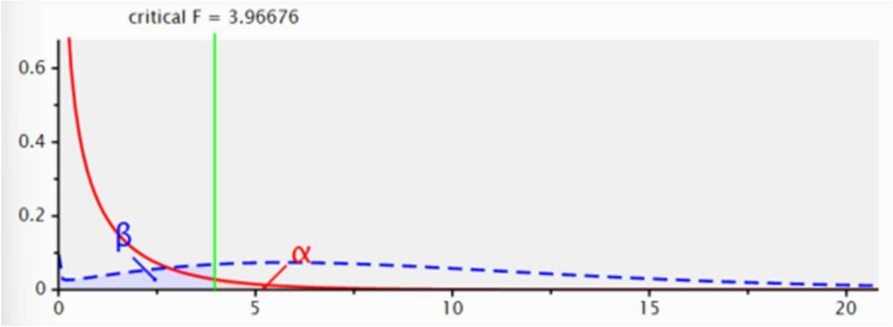

This configuration yielded: Total sample size = 78, matching the value reported in the article.

The power calculation reported by the study was based on a medium effect size (cf. Hedges' g = 0.531) derived from a previous meta-analysis (Konkolÿ Thege et al., 2021). However, the applicability of this effect size to the present RCT appears limited. The meta-analytic estimate pooled highly heterogeneous designs, including controlled and uncontrolled studies, clinical and non-clinical populations, and pre–post intervention effects (Leidy & Weissfeld, 1991; Matthay et al., 2021). Consequently, the reported g value primarily reflects aggregated within-group changes rather than between-group causal effects typically targeted in RCTs.

Because standardised pre–post effects are often larger than between-group differences, using this estimate as the basis for an a priori power calculation may have resulted in an optimistic assumption regarding the expected effect magnitude. Our reconstruction of the power analysis indicated that the reported sample size (N = 78) corresponds to a repeated measures ANOVA targeting the between-factors effect (f = 0.25), rather than the interaction effect implied by the textual description. Importantly, this calculation assumes balanced allocation, no attrition, and a moderate correlation (r ≈ 0.40) among repeated measures.

Furthermore, the final analysed sample (N = 68) fell below the planned sample size, suggesting that the achieved statistical power was likely lower than the intended 80%. These considerations indicate that the study may have been underpowered to detect the hypothesised effects. Underpowered designs increase the probability of Type II errors, yield unstable effect size estimates, and limit the interpretability of null or non-significant findings (Turner et al., 2013; Shreffler & Huecker, 2023).

Study 4 – Weinhold et al. (2013)

The sample size calculation reported by Weinhold et al. (2013) was independently replicated using G*Power version 3.1.9.7 to evaluate whether the parameters described in the manuscript allowed direct reproducibility.

The authors stated that an a priori power analysis was conducted for a repeated-measures mixed-design analysis of variance (ANOVA) using G*Power, indicating that a total sample size of 208 participants (104 per arm) would be sufficient to detect a small effect size (Cohen's f = 0.10) with **"a power of about 90%"** at an alpha level of .05.

1. Repeated measures within–between interaction

The first attempt used only parameters explicitly described in the manuscript:

- F tests → ANOVA: Repeated measures, within–between interaction
- A priori: Compute required sample size – given α, power, and effect size
- Effect size f = 0.10
- α = 0.05
- Power = 0.90
- Two groups
- Measurements = 3

**Figure A6.** Study 4 - central and noncentral distributions, first attempt

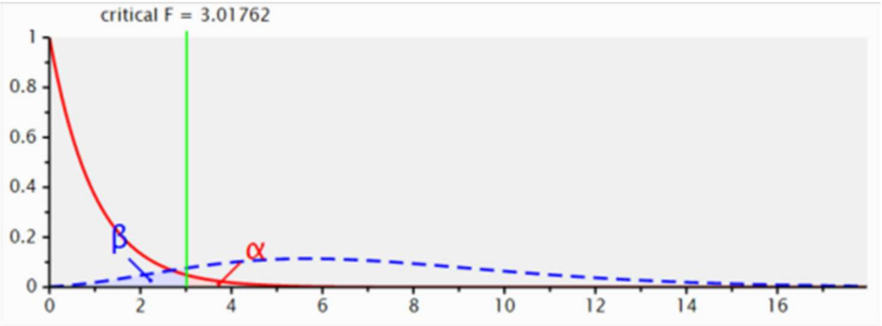

Because the manuscript did not report assumptions regarding the correlation among repeated measures or nonsphericity correction, default values commonly used in G*Power were applied (r = 0.50; ε = 1.00). This configuration yielded a required total sample size of N = 214, exceeding the reported N = 208.

This result led to the interpretation that the calculation could not be directly replicated using power = 0.90, suggesting that the phrase "about 90% power" did not correspond to an exact value of 0.90 in the software (statistical power).

2. Adjustment of correlation among repeated measures

Given the discrepancy, the correlation among repeated measures was incrementally increased ($r = 0.55$ and $r = 0.60$) while keeping power fixed at 0.90. These adjustments substantially reduced the required sample size ($N = 192$ and $N = 172$, respectively), indicating that correlation assumptions strongly influenced the output. These values deviated markedly from the reported $N = 208$, suggesting that correlation assumptions alone could not explain the discrepancy and the replication attempt had failed.

3. Adjustment of statistical power

Because the manuscript described power as "about 90%," additional attempts explored nearby values. Reducing power to 0.89 yielded $N = 206$, whereas specifying power = 0.895 produced $N = 210$. These results indicated that the reported sample size lay between these values.

4. Repeated measures ANOVA — within-between interaction

The reported sample size ($N = 208$) was reproduced when specifying:

- F tests → ANOVA: Repeated measures, within–between interaction
- A priori: Compute required sample size – given α, power, and effect size
- Effect size $f = 0.10$ (small)
- $\alpha = 0.05$
- Power = 0.892
- Two groups
- Measurements = 3
- Correlation among repeated measures $r = 0.50$
- Nonsphericity correction $\varepsilon = 1$

**Figure A7.** Study 4 - central and noncentral distributions, sixth attempt

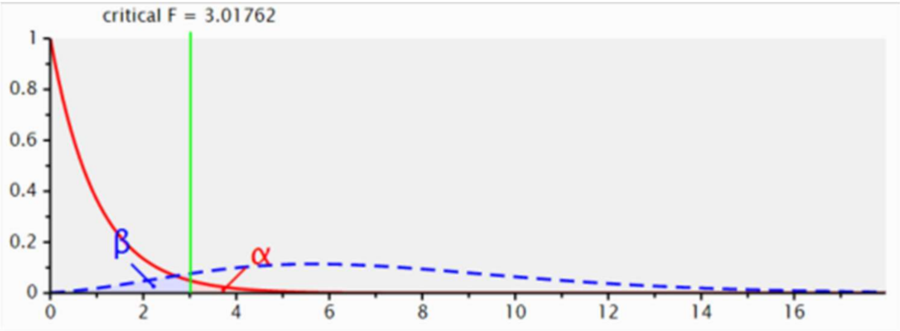

Under these assumptions, G*Power yielded N = 208, matching the value reported by the authors.

Although the trial reported using an intention-to-treat (ITT) framework, attrition remains relevant when considering statistical power. While ITT helps preserve the benefits of randomisation (McCoy, 2017), it does not fully compensate for the loss of information caused by missing follow-up data. In this study, the sample size planning did not include additional recruitment to account for attrition. Empirical evidence suggests that even modest withdrawals between randomisation and exposure can reduce effective power, particularly in trials targeting small effects (Kochovska et al., 2020).

The priori calculation indicated a nominal power of approximately 89%; however, attrition and unreported assumptions regarding the repeated-measures structure may have reduced the study's effective statistical precision. Given that the trial was powered to detect a small effect (f = 0.10), even relatively small losses of data can meaningfully influence the ability to detect differences over time. Although the available information does not allow a definitive conclusion that the study was underpowered, it is plausible that the achieved power was lower than originally anticipated.

Another concern regarding the sample size calculation reported by Weinhold and colleagues (2013) relates to the fact that the recruitment target of 104 participants per arm was divided between individuals actively receiving the intervention and those attending the session as "observers" of the constellation process. In clinical research, trials are typically designed to ensure that the analysed sample represents participants who are exposed to the intervention under evaluation, allowing the study to detect its effect relative to a comparison group. In this case, however, the intervention arm appears to include both treated participants and individuals who merely observed the session dynamics. This design raises questions about how the findings should be interpreted from a clinical perspective. If only a subset of participants actually received the therapeutic exposure, it becomes unclear whether any observed effects can be attributed to the intervention itself. Furthermore, this ambiguity complicates the translation of the results into clinical practice: when a patient seeks referral to a Family Constellation session, should clinicians recommend active participation in the constellation process or attendance as an observer?

Study 5 – NCT06863948

The ClinicalTrials registration (NCT06863948) describes the study as a pilot RCT primarily intended to inform methodology and effect size estimates for a future fully powered trial. While pilot studies may include clinical outcome measures, their primary function is to assess feasibility rather than to provide definitive evidence of efficacy (Leon et al., 2011; Eldridge et al., 2016). Notably, one of the stated aims is to evaluate reductions in anxiety and depressive symptoms compared to control conditions. Given that pilot trials are not powered to detect clinically meaningful effects, framing clinical outcome evaluation as a core objective may blur the distinction between exploratory and confirmatory research aims (Sim, 2019).

**Appendix 12 -** Characteristics of trials included in the review

**Table A12.** Characteristics extracted from Goode (2016)

| | |
|---|---|
| Article ID: | 1 |
| Article title: | Enhancing the affective domain in order to reduce fear of death in first-year student nurses |
| Article link: | https://uhra.herts.ac.uk/id/eprint/16098/ |
| Notes ("N/A" if not applicable): | Unpublished / Not peer-reviewed / Grey literature |
| Firs author (last name): | Goode |

| **Study characteristics** | | | | | | |
|---|---|---|---|---|---|---|
| Country of correspondence author | Country where study conducted | Language of publication | Study design | Funding source | Dates study began and ended | Number of centres |
| England | England | English | Randomised controlled trial | No funding | 2011-2013 | 1 |

| **Study characteristics** | | | |
|---|---|---|---|
| Ethical approval obtained (yes/no?) | Conflicts of interest | Journal published | Impact factor |
| Yes | Not reported / The author is a FCT facilitator | Not published | NA |

| **Characteristics of Participants** | | | | | | |
|---|---|---|---|---|---|---|
| Detailed population description (e.g., diagnostic status) | Diagnosis (if applicable and how it was established) | Setting | Inclusion criteria | Exclusion criteria | Method of recruitment of participants | Informed consent obtained (yes/no)? |
| First-semester student nurses (Non-clinical sample) | NA | Non-clinical sample | Not reported | Not reported | In-person recruitment of the entire February 2011 adult student cohort via verbal presentation by the researcher; voluntary participation following study explanation | Yes |

| **Characteristics of Participants** | | | | | |
|---|---|---|---|---|---|
| Total number of assessed for eligibility | Total number recruited | Total number randomised (sample size) | Total number of withdrawals, drop-outs and exclusions | Clusters (number, type, number of people per cluster) | Baseline imbalances (describe in detail if applicable) |
| ~100 students (entire February 2011 adult student cohort) | 76 | 75 | 20 | NA | Age and previous experience were reported by group only at stage 2 (8-months), precluding assessment of baseline comparability |

| Characteristics of Participants | | | |
|---|---|---|---|
| Age (mean + SD, or median + IQR; make sure to distinguish between IQR and RANGE) | Sex (and proportions, %) | Race/ethnicity (and proportions, %) | Co-morbidities |
| N = 75: 18–20: 19 (25%); 21–25: 20 (27%); 26–30: 11 (15%); 31–36: 6 (8%); >36: 19 (25%) | Male: 3/75 (4%) | White 40 (53%); Mixed/Multiple 6 (8%); Asian/Asian British 5 (7%); Black/African/Caribbean/Black British 24 (32%) | NA |

| Characteristics of interventions | | | | | | |
|---|---|---|---|---|---|---|
| Group name | Number randomised to group | Demographics of group (age, race/ethnicity, sex) | Description (e.g., content, dose, components) | Duration of treatment period | Timing (e.g., frequency, duration) | Delivery (e.g., mechanism, medium, intensity, fidelity) |
| Do Something Different (DSD) | 25 | Baseline data not reported separately by group | Participants received a copy of the DSD book and were instructed to dedicate a few minutes each day to completing structured exercises from the book | 6 weeks | Daily | Self-applied |
| Family Constellations (FC) | 25 | Baseline data not reported separately by group | Group intervention using Family Constellations, including reflective sessions, symbolic representation of family members, exploration of relational issues, discussion, and final reflection at the end of each session | 6 weeks | Weekly, 1 hour per session | In-person, group intervention |
| Control | 25 | Baseline data not reported separately by group | No intervention | NA | NA | NA |

| Characteristics of interventions | | | | | |
|---|---|---|---|---|---|
| Providers (e.g., no profession, training, ethnicity etc., if relevant) | Co-interventions | Integrity of delivery | Compliance | Family involvement | Concurrent treatments? |

| | | | | | |
|---|---|---|---|---|---|
| Self-applied | Not reported | NA | Not assessed. Adherence to daily DSD exercises was encouraged but not formally measured.<br><br>“It was not possible to assess how many of the 25 students completed this task”<br><br>“Difficult to discern to what extent the DSD had been completed” | NA | Not reported |
| Sessions facilitated by the researcher. The researcher had completed two years of formal training in Family Constellations before the start of the program | Not reported | NA | Only 14/25 participants (56%) attended at least one session; 7/25 (28%) attended all six sessions | NA | Not reported |
| NA | Not reported | NA | NA | NA | Not reported |

| **Outcomes (for each outcome, including side-effects)** | | | | | | |
|---|---|---|---|---|---|---|
| Outcome name | Measurement tool(s) | Time points measured | Time points reported | Outcome definition | Person measuring/ reporting | Unit of measurement (will generally be absolute points within a scale) |
| Multidimensional Fear of Death Scales | Multidimensional Fear of Death Scale (MFODS) | Baseline; 8-months | Baseline; 8-months | Fear of death and dying | Self-reported by participants using a questionnaire | Absolute points on a 5-point Likert scale (MFODS total and/or subscale scores) |
| Adverse effects | NA | Not reported | Not reported | NA | NA | NA |

| Upper and lower limits (indicate whether high or low score is good) | Imputation of missing data (e.g. assumptions made for ITT analysis) | Power (e.g. power & sample size calculation) |
|---|---|---|
| 1–5 per item; higher scores indicate greater fear of death (higher is worse) | NA | No reported |
| NA | NA | NA |

**Table A13.** Characteristics extracted from Hunger et al. (2014)

| | |
|---|---|
| Article ID: | 2 |
| Article title: | Improving Experience in Personal Social Systems through Family Constellation Seminars: Results of a Randomized Controlled Trial |
| Article link: | https://doi.org/10.1111/famp.12051 |
| Notes ("N/A" if not applicable): | This RCT is the same as ID #4 |
| Firs author (last name): | Hunger |

| **Study characteristics** | | | | | | |
|---|---|---|---|---|---|---|
| Country of correspondence author | Country where study conducted | Language of publication | Study design | Funding source | Dates study began and ended | Number of centres |
| Germany | Germany | English | Randomised controlled trial | DFG (German Research Foundation) | Fev/2011 to Dec/2011 | 1 |

| **Study characteristics** | | | |
|---|---|---|---|
| Ethical approval obtained (yes/no?) | Conflicts of interest | Journal published | Impact factor |
| Yes | Not reported | Family Process | 2.2 |

| **Characteristics of Participants** | | | | | | |
|---|---|---|---|---|---|---|
| Detailed population description (e.g., diagnostic status) | Diagnosis (if applicable and how it was established) | Setting | Inclusion criteria | Exclusion criteria | Method of recruitment of participants | Informed consent obtained (yes/no)? |
| Adults (≥18 years) participating in Family Constellation Seminars; participants did not need a clinical diagnosis or to be in treatment; participants could be in counselling and/or psychotherapy; included both “active participants” (presenting their own constellation) and “observing participants” (representatives only) | No clinical diagnosis required; no diagnostic assessment procedure reported | Outpatients | (a) Active participants had to sign up for a family constellation of their own and present a serious conflictual social systems dynamic they wanted to work with; (b) Observing participants had to have an interest in participating in a FCS but as representatives only; (c) All participants had to participate in no further FCS until completion of the study; (d) Minimum age of 18 years | Not reported | Information flyer and a website linked to institutes and associations of systemic counseling and psychotherapy; eligibility assessed via telephone interviews | Yes |

| Total number of assessed for eligibility | Total number recruited | Total number randomised (sample size) | Total number of withdrawals, drop-outs and exclusions | Clusters (number, type, number of people per cluster) | Baseline imbalances (describe in detail if applicable) | Age (mean + SD, or median + IQR; make sure to distinguish between IQR and RANGE) | Sex (and proportions, %) | Race/ethnicity (and proportions, %) | Co-morbidities |
|---|---|---|---|---|---|---|---|---|---|
| 371 | 208 | 208 | 17 | NA | There are more participants married or living with a partner in the control group (8% difference) | Intervention group: 47 (9); Wait-list group: 48 (10) | Intervention group: Female 87/104 (84%); Wait-list group: Female 77/104 (74%) | German nationality: Intervention group 101/104 (97%); Wait-list control group 99/104 (95%). No other race/ethnicity categories reported | Not reported |

**Characteristics of interventions**

| Group name | Number randomised to group | Demographics of group (age, race/ethnicity, sex) | Description (e.g., content, dose, components) | Duration of treatment period | Timing (e.g., frequency, duration) | Delivery (e.g., mechanism, medium, intensity, fidelity) | Providers (e.g., no profession, training, ethnicity etc., if relevant) | Co-interventions | Integrity of delivery | Compliance |
|---|---|---|---|---|---|---|---|---|---|---|
| Family Constellation Seminars | 104 | Age: 47 (9); Female: 87/104 (84%); Married or living with a partner 69(66%); High-school diploma 92(89%); Employed: 98 (96%); German nationality: 101/104 (97%); Previous experience with FCS: 83(80%) | Group-based intervention including active and observing participants, involving problem and solution constellations with facilitator-led procedures. Obligatory items: (a) welcome and information; (b) explanation of general philosophy and FCS procedures; (c) explanation of facilitator's working style; (d) instructions for representatives and how to communicate from that role; (e) instructions on how to position representatives in the room; (f) time for participant questions; and (g) an initial round to clarify active participants' goals | 3-day seminar | Nonrecurring | In-person, group-based seminar | Two facilitators: one male licensed psychiatrist and psychotherapist (30 years' experience); one female clinical psychologist and licensed psychotherapist (20 years' experience) | NA | NA | 99/104 participants in intervention group received FCS; 5 did not |

| | | | | | | | | | | |
|---|---|---|---|---|---|---|---|---|---|---|
| Waitlist | 104 | Age: 48 (10); Female: 77/104 (74%); Married or living with a partner 77(74%); High-school diploma 89(86%); Employed: 94(90%); German nationality: 99/104 (95%); Previous experience with FCS: 82(79%) | NA | 4 months | NA | NA | NA | NA | NA | 4 participated in another FCS before completion of study |

| **Characteristics of interventions** | |
|---|---|
| Family involvement | Concurrent treatments? |
| NA | Participants were allowed to be in counseling and/or psychotherapy |
| Not reported | Participants were allowed to be in counseling and/or psychotherapy |

| **Outcomes (for each outcome, including side-effects)** | | | | | | | | | |
|---|---|---|---|---|---|---|---|---|---|
| Outcome name | Measurement tool(s) | Time points measured | Time points reported | Outcome definition | Person measuring/ reporting | Unit of measurement (will generally be absolute points within a scale) | Upper and lower limits (indicate whether high or low score is good) | Imputation of missing data (e.g. assumptions made for ITT analysis) | Power (e.g. power & sample size calculation) |

| | | | | | | | | | |
|---|---|---|---|---|---|---|---|---|---|
| Experience in personal social systems | Experience In Social Systems Questionnaire, personal domain (EXIS.pers) | Baseline, 2 weeks, 4 months | Baseline, 2 weeks, 4 months | Participants' experience within their personal social systems, reflecting belonging, autonomy, accord, and confidence over the previous 2 weeks | Self-reported by participants using a questionnaire | Absolute points on a Likert-type scale; 12 items rated from 1 to 6; total score derived from item ratings | Lower limit: 1 ("not at all"); Upper limit: 6 ("entirely"); (lower is worse) | Single missing values for each (sub-)scale were replaced with conditional mean values calculated within predefined subgroups (active/observing × intervention/wait-list) | Not reported |
| Adverse effects | Passive surveillance | Entire intervention period up to 4-month follow-up | Entire intervention period up to 4-month follow-up | Occurrence of adverse events reported by participants | Self-report | Count / presence of adverse events | NA | NA | NA |

**Table A14.** Characteristics extracted from Thege & Szabó (2024)

| | |
|---|---|
| Article ID: | 3 |
| Article title: | The efficacy of pandemic-adjusted family/systemic constellation therapy in improving psychopathological symptoms: A randomized controlled trial |
| Article link: | https://doi.org/10.1016/j.jpsychires.2024.07.027 |
| Notes ("N/A" if not applicable): | N/A |
| Firs author (last name): | Thege |

| **Study characteristics** | | | | | | |
|---|---|---|---|---|---|---|
| Country of correspondence author | Country where study conducted | Language of publication | Study design | Funding source | Dates study began and ended | Number of centres |
| Canada | Hungary | English | Randomised controlled trial | Károli Gáspár University of the Reformed Church in Hungary | 19/07/2021 to 25/4/2022 | 1 |

| **Study characteristics** | | | |
|---|---|---|---|
| Ethical approval obtained (yes/no?) | Conflicts of interest | Journal published | Impact factor |
| Yes | No | Journal of Psychiatric Research | 3.2 |

| **Characteristics of Participants** | | | | | | |
|---|---|---|---|---|---|---|
| Detailed population description (e.g., diagnostic status) | Diagnosis (if applicable and how it was established) | Setting | Inclusion criteria | Exclusion criteria | Method of recruitment of participants | Informed consent obtained (yes/no)? |
| Adults (≥18 years) recruited from the general population in Hungary; participants willing to attend a family/systemic constellation seminar; participants without a current mental disorder diagnosis | No diagnosis required for inclusion; exclusion applied if there was a current mental disorder diagnosed by a health care professional | Outpatients | (a) At least 18 years old; (b) Willingness to participate in a family/systemic constellation seminar on a date determined by randomisation, with no other date during the study period. | (a) Current mental disorder diagnosed by a health care professional; (b) Recent (<1 year) prior participation in a family/systemic constellation seminar in another setting | Paid social media advertisements targeting the capital city in Hungary | Yes |

| **Characteristics of Participants** | | | | | | | | | |
|---|---|---|---|---|---|---|---|---|---|
| Total number of assessed for eligibility | Total number recruited | Total number randomised (sample size) | Total number of withdrawals, drop-outs and exclusions | Clusters (number, type, number of people per cluster) | Baseline imbalances (describe in detail if applicable) | Age (mean + SD, or median + IQR; make sure to distinguish between IQR and RANGE) | Sex (and proportions, %) | Race/ethnicity (and proportions, %) | Co-morbidities |

| | | | | | | | | | |
|---|---|---|---|---|---|---|---|---|---|
| 80 | 80 | 80 | 12 | NA | A statistically significant difference between groups was reported for prior experience with systemic constellations | Total sample: 41.9 (9.2) years; Intervention group: 42.2 (8.2) years; Control group: 41.6 (10.4) years | Total sample: Men 22 (32.4%); Women 46 (67.6%); Intervention group: Men 12 (34.3%); Women 23 (65.7%); Control group: Men 10 (30.3%); Women 23 (69.7%) | Not reported | Not reported |

| **Characteristics of interventions** | | | | | | |
|---|---|---|---|---|---|---|
| Group name | Number randomised to group | Demographics of group (age, race/ethnicity, sex) | Description (e.g., content, dose, components) | Duration of treatment period | Timing (e.g., frequency, duration) | Delivery (e.g., mechanism, medium, intensity, fidelity) |
| Intervention group | 40 | Age: 42.2 (8.2); Men 12 (34.3%); Women 23 (65.7%); Educational attainment - Secondary: 8 (22.9); Educational attainment - Post-secondary: 27 (77.1); Marital status - Single: 8 (22.9); Marital status - In relationship: 21 (60.0); Marital status - Separated/divorced: 6 (17.1); Prior experience with systemic constellations: 16 (45.7) | Family/systemic constellation therapy delivered in a group setting, including: brief interview between the active client ("issue holder") and facilitator; selection of system members/elements; representation by other group members; spatial positioning; non-verbal experiential focus; inquiry into representatives' sensations, feelings, and thoughts; rearrangements, spatial adjustments, and ritualized conversations; development of a "solution constellation" | Single-day workshops (pandemic-adjusted), instead of the traditional two-to-three-day format | One-time participation in a single-day workshop; four single-day workshops were conducted in total | In-person, group-based delivery; approximately 10 participants per workshop; mask wearing and physical distancing required; adherence to a previously developed intervention protocol assessed by independent observers; high treatment integrity reported for mandatory and optional procedures |
| Waitlist control group | 40 | Age: 41.6 (10.4); Men 10 (30.3%); Women 23 (69.7%); Educational attainment - Secondary: 11 (33.3); Educational attainment - Post-secondary: 22 (66.7); Marital status - Single: 6 (18.2); Marital status - In relationship: 22 (66.7); Marital status - Separated/divorced: 5 (15.2); Prior experience with systemic constellations: 7 (21.2) | NA | 6 months | NA | NA |

| **Characteristics of interventions** | | | | | |
|---|---|---|---|---|---|
| Providers (e.g., no profession, training, ethnicity etc., if relevant) | Co-interventions | Integrity of delivery | Compliance | Family involvement | Concurrent treatments? |

| | | | | | |
|---|---|---|---|---|---|
| Two intervention providers: one clinical psychologist and one physician; highly trained in general mental health care delivery; 10–20+ years of experience in family constellation therapy; both led two single-day workshops | NA | NA | Very high adherence to the intervention protocol both in terms of the mandatory (98.8–100% depending on workshop day) and optional (84.1–100% across the four workshop days) procedures | NA | NA |
| NA | NA | NA | Two participants were excluded due protocol deviation (participating in a family constellation workshop in another setting between the T2 and T3) | NA | NA |

| **Outcomes (for each outcome, including side-effects)** | | | | | |
|---|---|---|---|---|---|
| Outcome name | Measurement tool(s) | Time points measured | Time points reported | Outcome definition | Person measuring/ reporting |
| Overall psychopathology | Brief Symptom Inventory (BSI) | Baseline; 1 month; 6 months | Baseline; 1 month; 6 months | Individual's overall psychopathology level in symptom dimensions: Somatization, Obsessive-compulsive symptoms, Interpersonal sensitivity, Depression, Anxiety, Hostility, Phobic anxiety, Paranoid ideation, Psychoticism | Self-reported by participants using a questionnaire |
| Adverse effects | Qualitative ad hoc questions | 1 month; 6 months | 1 month; 6 months | Perceived negative effects related to participation in the intervention. | Self-reported by participants using two qualitative questions |

| **Outcomes (for each outcome, including side-effects)** | | | |
|---|---|---|---|
| Unit of measurement (will generally be absolute points within a scale) | Upper and lower limits (indicate whether high or low score is good) | Imputation of missing data (e.g. assumptions made for ITT analysis) | Power (e.g. power & sample size calculation) |

| | | | |
|---|---|---|---|
| Absolute points on a scale | Not reported | NA | A priori power analysis (GPower): repeated-measures mixed-design ANOVA; medium effect (Hedges' g = 0.531); 80% power; $\alpha = 0.05$; required N = 78 (39 per group) |
| Count / presence of adverse events | NA | NA | NA |

**Table A15.** Characteristics extracted from Weinhold (2013)

| | |
|---|---|
| Article ID: | 4 |
| Article title: | Family Constellation Seminars Improve Psychological Functioning in a General Population Sample: Results of a Randomized Controlled Trial |
| Article link: | https://doi.org/10.1037/a0033539 |
| Notes ("N/A" if not applicable): | This RCT is the same as ID #2 |
| Firs author (last name): | Weinhold |

| Study characteristics | | | | | | |
|---|---|---|---|---|---|---|
| Country of correspondence author | Country where study conducted | Language of publication | Study design | Funding source | Dates study began and ended | Number of centres |
| Germany | Germany | English | Randomised controlled trial | DFG (German Research Foundation) | Fev/2011 to Dec/2011 | 1 |

| Study characteristics | | | |
|---|---|---|---|
| Ethical approval obtained (yes/no?) | Conflicts of interest | Journal published | Impact factor |
| Yes | Not reported | Journal of Counseling Psychology | 3.8 |

| Characteristics of Participants | | | | | | |
|---|---|---|---|---|---|---|
| Detailed population description (e.g., diagnostic status) | Diagnosis (if applicable and how it was established) | Setting | Inclusion criteria | Exclusion criteria | Method of recruitment of participants | Informed consent obtained (yes/no)? |
| Adults (≥18 years) participating in Family Constellation Seminars; participants did not need a clinical diagnosis or to be in treatment; participants could be in counseling and/or psychotherapy; included both “active participants” (presenting their own constellation) and “observing participants” (representatives only) | No clinical diagnosis required; no diagnostic assessment procedure reported | Outpatients | (a) Active participants had to sign up for a family constellation of their own and present a serious conflictual social systems dynamic they wanted to work with; (b) Observing participants had to have an interest in participating in a FCS but as representatives only; (c) All participants had to participate in no further FCS until completion of the study; (d) Minimum age of 18 years | Not reported | Information flyer and a website linked to institutes and associations of systemic counseling and psychotherapy; eligibility assessed via telephone interviews | Yes |

| Characteristics of Participants |
|---|

| Total number of assessed for eligibility | Total number recruited | Total number randomised (sample size) | Total number of withdrawals, drop-outs and exclusions | Clusters (number, type, number of people per cluster) | Baseline imbalances (describe in detail if applicable) | Age (mean + SD, or median + IQR; make sure to distinguish between IQR and RANGE) |
|---|---|---|---|---|---|---|
| 371 | 208 | 208 | 15 | NA | There are more participants married or living with a partner in the control group (8% difference) | Intervention group: 47 (9); Wait-list group: 48 (10) |

**Characteristics of Participants**

| Sex (and proportions, %) | Race/ethnicity (and proportions, %) | Co-morbidities |
|---|---|---|
| Intervention group: Female 87/104 (84%); Wait-list group: Female 77/104 (74%) | German nationality: Intervention group 101/104 (97%); Wait-list control group 99/104 (95%). No other race/ethnicity categories reported | Not reported |

**Characteristics of interventions**

| Group name | Number randomised to group | Demographics of group (age, race/ethnicity, sex) | Description (e.g., content, dose, components) | Duration of treatment period | Timing (e.g., frequency, duration) | Delivery (e.g., mechanism, medium, intensity, fidelity) |
|---|---|---|---|---|---|---|
| Family Constellation Seminars | 104 | Age: 47 (9); Female: 87/104 (84%); Married or living with a partner 69(66%); High-school diploma 92(89%); Employed: 98 (96%); German nationality: 101/104 (97%); Previous experience with FCS: 83(80%) | Group-based intervention including active and observing participants, involving problem and solution constellations with facilitator-led procedures. Obligatory items: (a) welcome and information; (b) explanation of general philosophy and FCS procedures; (c) explanation of facilitator’s working style; (d) instructions for representatives and how to communicate from that role; (e) instructions on how to position representatives in the room; (f) time for participant questions; and (g) an initial round to clarify active participants’ goals | 3-day seminar | Nonrecurring | In-person, group-based seminar |

| | | | | | | |
|---|---|---|---|---|---|---|
| Waitlist | 104 | Age: 48 (10); Female: 77/104 (74%); Married or living with a partner 77(74%); High-school diploma 89(86%); Employed: 94(90%); German nationality: 99/104 (95%); Previous experience with FCS: 82(79%) | NA | 4 months | NA | NA |

| **Characteristics of interventions** | | | | | |
|---|---|---|---|---|---|
| Providers (e.g., no profession, training, ethnicity etc., if relevant) | Co-interventions | Integrity of delivery | Compliance | Family involvement | Concurrent treatments? |
| Two facilitators: one male licensed psychiatrist and psychotherapist (30 years' experience); one female clinical psychologist and licensed psychotherapist (20 years' experience) | NA | NA | 99/104 participants in intervention group received FCS; 5 did not | NA | Participants were allowed to be in counseling and/or psychotherapy |
| NA | NA | NA | 4 participated in another FCS before completion of study | Not reported | Participants were allowed to be in counseling and/or psychotherapy |

| **Outcomes (for each outcome, including side-effects)** | | | | | | |
|---|---|---|---|---|---|---|
| Outcome name | Measurement tool(s) | Time points measured | Time points reported | Outcome definition | Person measuring/ reporting | Unit of measurement (will generally be absolute points within a scale) |
| Psychological functioning | The Outcome Questionnaire (OQ-45.2) | Baseline, 2 weeks, 4 months | Baseline, 2 weeks, 4 months | Psychotherapeutic change over the previous week | Self-reported by participants using a questionnaire | Absolute points on OQ-45.2 total score (TOT) |

| | | | | | | |
|---|---|---|---|---|---|---|
| Adverse effects | Passive surveillance | Entire intervention period up to 4-month follow-up | Entire intervention period up to 4-month follow-up | Occurrence of adverse events reported by participants | Self-report | Count / presence of adverse events |

| **Outcomes (for each outcome, including side-effects)** | | |
|---|---|---|
| Upper and lower limits (indicate whether high or low score is good) | Imputation of missing data (e.g. assumptions made for ITT analysis) | Power (e.g. power & sample size calculation) |
| Upper and Lower limits not reported; Outcome direction: higher is worse | Yes; Single missing values <20% replaced with conditional mean values | A priori power analysis (G*Power): repeated-measures mixed-design ANOVA; small effect (Cohen's f = 0.1); ~90% power; α = 0.05; required N = 208 (104 per group). |
| NA | NA | NA |
